# Risk factors for asthma-related hospital and intensive care admissions in children, adolescents, and adults: a cohort study using primary and secondary care data

**DOI:** 10.1101/2022.11.11.22282223

**Authors:** Nikita Simms-Williams, Prasad Nagakumar, Rasiah Thayakaran, Nicola J Adderley, Richard Hotham, Adel H Mansur, Krishnarajah Nirantharakumar, Shamil Haroon

## Abstract

**Objectives:** To assess the association between demographic and clinical risk factors and asthma-related hospital and intensive care admissions in children, adolescents, and adults, and to estimate the proportion of hospital admissions attributable to modifiable risk factors.

**Design:** Cohort study using routinely collected primary and secondary care data.

**Setting:** A large UK-based primary care database, the Clinical Practice Research Datalink (CPRD) Aurum, and linked Hospital Episode Statistics Admitted Patient Care (HES APC) data.

**Participants:** Patients were eligible for the study if they were aged five years and older and had an asthma diagnosis with linked data to the HES APC database. This included 90,989 children aged 5-11 years, 114,927 adolescents aged 12-17 years, and 1,179,410 adults aged 18 years or older.

**Primary and secondary outcome measures:** Primary outcome: asthma-related hospital admissions recorded from 1^st^ January 2017 to 31^st^ December 2019. Secondary outcome: asthma-related intensive care unit (ICU) admissions. Incidence rate ratios (IRR) adjusted for demographic and clinical risk factors were estimated using negative binomial models. Population attributable fraction (PAF) amongst those with asthma was estimated for modifiable risk factors that were statistically significantly associated with the primary outcome.

**Results:** In children, the risk factors for asthma-related hospital admission were belonging to an ethnic minority group, increasing socioeconomic deprivation, allergies (PAF 11.4%, 95% CI 6.8 to 15.8), and atopic eczema (6.8%, 3.6 to 9.9). In adolescents, the risk factors were being female, belonging to an ethnic minority group, increasing socioeconomic deprivation, former smoking (PAF 6.8%, 0.9 to 12.3), and allergic rhinitis. In adults, the risk factors were younger age, being female, belong to an ethnic minority group, increasing socioeconomic deprivation, being underweight, overweight or obese (PAF 23.3%, 95% CI 20.5 to 26.1 for obesity), current smoking (4.3%, 3.0 to 5.7), depression (11.1%, 9.1 to 13.1), allergies (6.2%, 4.4 to 8.0), gastro-oesophageal reflux disease (2.3%, 1.2 to 3.4), anxiety (2.0%, 0.5 to 3.6), and chronic rhinosinusitis (0.8%, 0.3 to 1.3%). In all age groups, increasing medication burden was associated with an increased risk in the primary outcome.

Risk factors for asthma-related ICU admissions in children were black or mixed ethnicity and high levels of socioeconomic deprivation; in adolescents, they were female sex and black ethnicity; and in adults, they were younger age, female sex, black, mixed, or Asian ethnicity, and depression. In all age groups, increasing medication burden was associated with an increased risk in the secondary outcome.

**Conclusions:** There are significant sociodemographic inequalities in the rates of asthma-related hospital and ICU admissions. Treating atopic conditions in all age groups should be considered an integral part of asthma management. Adults have a wide range of potentially treatable risk factors that contribute substantially to asthma-related hospital admissions, including obesity, smoking, depression, anxiety and gastro-oesophageal reflux disease. Treating these risk factors could significantly reduce the rate of avoidable hospital admissions. Overall asthma medication burden is an important reflection of disease severity and prognostic marker of asthma outcomes, which should be monitored in all patients.

**WHAT IS ALREADY KNOWN ON THIS TOPIC:**

- Asthma is one of the most common chronic diseases and remains an important cause of avoidable hospital and intensive care admissions.
- Risk factors for asthma have previously been described but there are a lack of large population scale analyses stratifying these risk factors among children, adolescents, and adults, or providing estimates of the key modifiable risk factors that most contribute to avoidable hospital admissions.

**WHAT THIS STUDY ADDS:**

- There are significant sociodemographic inequalities in asthma-related hospital and intensive care admissions in children, adolescents, and adults.
- Atopic disorders and smoking are key addressable risk factors in all age groups, while obesity, depression, and anxiety are important treatment targets more specific to adults.
- Overall asthma medication burden is strongly associated with the risk of asthma-related hospital and ICU admissions and should be used for assessing disease severity and monitoring asthma control and prognosis.

## INTRODUCTION

Asthma is the most common chronic respiratory disease affecting children and adults with an estimated worldwide prevalence of 10% and affecting over 600 million people in 2019.^1^ The UK has one of the highest prevalence of asthma and associated morbidity and mortality in western Europe, accounting for over 1,000 deaths and 60,000 emergency hospital admissions per year. ^2 3^ This has resulted in a significant and preventable health and economic burden that accounts for 200,000 bed days per year and costs the UK National Health Service £1.1 billion per year.^3^ Despite efforts to improve the management of asthma through implementation of national clinical guidelines,^4^ and publication of the National Asthma Death Review (NRAD) recommendations,^5^ there has been little reduction in asthma attacks across all age groups in the UK.^6^

Asthma attacks severe enough to require hospital admission often indicate poor management of modifiable risk factors, including comorbidities.^7 8^ Several risk factors for asthma attacks have been described separately in children and adults including female sex,^9 10 11 12^ a history of previous asthma-related hospital admissions,^10 13^ higher disease severity,^14^ older age,^9 12^ belonging to an ethnic minority group,^13 15^having a history of inhaled corticosteroid use,^15^ and presence of comorbidities such as allergic rhinitis, gastro-oesophageal reflux disease and obesity.^11 16^ However, risk factors contributing to asthma attacks have not been studied across paediatric and adult populations simultaneously in a large population representative sample. Furthermore, asthma management guidelines advise stepping up medications to improve control but there is relatively little emphasis on addressing modifiable risk factors and comorbidities at each step of the treatment pathway, highlighting the need for more research focusing on their contribution to the burden of asthma exacerbations.

Although risk factors for severe asthma attacks have been described in children and adults,^17^ these are likely to be different in primary and secondary school age children and across adult age groups. Furthermore, the impact of addressing modifiable risk factors, including comorbidities, on severe asthma attacks across different age groups is also unknown. There is therefore a need to characterise risk factors and comorbidities unique to different age groups using large population representative datasets to inform the development of interventions aimed at reducing asthma related hospitalisations and healthcare costs and improving quality of life.

The aim of the study was to investigate risks factors associated with asthma-related hospitalisations and intensive care unit (ICU) admissions. The specific objectives were to estimate the incidence rates of asthma-related hospital and intensive care admissions in children, adolescents, and adults, estimate the association between demographic and clinical risk factors and these outcomes, and estimate the proportion of hospital admissions that could be prevented by addressing key modifiable risk factors.

## METHODS

### Study design

This was a cohort study using routinely collected primary and secondary care data to assess the incidence of asthma-related hospital and intensive care unit (ICU) admissions in different age groups, the association between demographic and clinical risk factors with these outcomes, and the proportion of hospital admissions that are theoretically preventable by addressing key modifiable risk factors.

### Setting and data source

UK primary care records were extracted from the Clinical Practice Research Datalink (CPRD) Aurum database with linked Hospital Episode Statistics (HES) Admitted Patient Care (APC) data^18^ for the period from 1^st^ January 2017 to 31^st^ December 2019. General practices contributing to CPRD Aurum cover approximately 19% of the UK population, all of which use the EMIS clinical information system which uses SNOMED-CT terms to code clinical data, and drug codes linked to the British National Formulary to code drug prescriptions.^19^ CPRD Aurum holds data for over 13 million currently registered patients.^20^ Over 90% of the data held within CPRD Aurum is linked to the HES database, which codes diagnoses using ICD-10 codes.^21^

### Population

General practices that had contributed data to CPRD Aurum for at least one year prior to the index date (1^st^ January 2017) were eligible for inclusion in the study. Data were extracted for patients aged five years and older with a diagnosis of asthma prior to the index date who had been registered with an eligible general practice for at least one year prior to the index date. Asthma was defined as the presence of a SNOMED-CT code for asthma, as listed in Supplementary table 1. The SNOMED-CT terms were selected using a systematic process with clinical input that involved checking existing code lists used by our research team, checking published code lists, searching the SNOMED-CT terminology browser, and searching using free text terms for asthma within an inhouse software tool called Code Builder.

Patients were excluded from the study if they had a diagnosis of chronic obstructive pulmonary disease (COPD), bronchiectasis, obstructive sleep apnoea or interstitial lung disease due to the significant potential for misclassification of the primary diagnosis for hospitalisation (e.g., hospitalisations for asthma being misclassified as being due to COPD). All patients were followed up from the index date (1^st^ January 2017) and follow-up was terminated at the earliest of death, exit from the database (patient left the practice or the practice stopped contributing to the database), or the study end date (31^st^ December 2019).

### Outcomes

The primary outcome was hospital admissions for asthma, assessed using linked secondary care (HES APC) data. Asthma admissions were defined as hospital admissions with associated ICD-10 asthma diagnosis codes J45 and J46 as the primary diagnostic code for the admission. The secondary outcome was asthma-related intensive care unit (ICU) admissions.

### Demographic and clinical risk factors

The baseline patient demographic characteristics and behavioural risk factors extracted prior to the index date included age, sex, ethnic group, socioeconomic status (measured using the Index of Multiple Deprivation quintile [IMD]),^22^ body mass index (BMI), and smoking status. The comorbidities extracted included atopic diseases (allergies, atopic dermatitis and allergic rhinitis), gastro-oesophageal reflux disease (GORD), chronic rhinosinusitis, and mental health disorders (anxiety and depression). Asthma-related prescriptions within one year prior to the index date were also extracted including short-acting bronchodilators (SABA), oral corticosteroids (OCS), inhaled corticosteroids (ICS), long-acting beta-2 agonists (LABA), long-acting muscarinic antagonists (LAMA), leukotriene receptor antagonists (LTRA), and influenza vaccination.

### Quantitative variables

Variables were categorised into the following groupings: age (5-11, 12-17, 18-24, 25-39, 40-59, 60-79 and ≥80 years), sex (male and female), ethnicity (white, black, mixed, Asian, other and missing), IMD score quintile (1 [least deprived], 2, 3, 4, 5 [most deprived] and missing), BMI (underweight [<18.5 kg/m^2^], normal weight [18.5-24], overweight [25-29], obese [≥30] and missing), smoking status (current smoker, former smoker, never smoked and missing), and number of SABA prescriptions (0, 1-3, 4-6 and >6). Standardised BMI measures were calculated using z scores for children and adolescents.

### Statistical methods

The cohort was stratified into children aged 5 to 11 years, adolescents aged 12 to 17 years, and adults aged 18 years and older. Descriptive statistics were used to describe the characteristics of the cohort including age, sex, ethnic group, socioeconomic status, smoking status, BMI, comorbidities, and prescriptions. Baseline data were described using frequencies and proportions for categorical variables and median and interquartile range (IQR) for continuous data with a skewed distribution. The incidence rates for asthma-related hospital and ICU admissions were calculated, stratified by age group.

Akaike’s Information Criteria (AIC) and Bayesian Information Criteria (BIC) values were generated for three different statistical models (Poisson regression, zero-inflated Poisson regression and negative binomial regression) to identify the most appropriate model for the analysis. Negative binomial regression was used instead of a Poisson regression model as the outcome of count data was over-dispersed (see appendix 1).

The covariates used in the analyses were as follows: demographic characteristics (age, sex, ethnic group, socioeconomic status, BMI, and smoking status), comorbidities (allergies, atopic dermatitis allergic rhinitis, GORD, chronic rhinosinusitis, anxiety, and depression), asthma-related prescriptions (SABA, OCS, ICS, LAMA, LABA and LTRA), and influenza vaccination. Separate negative binomial regression models were used for the three age groups to assess the association between the pre-specified covariates and the primary and secondary outcomes, presented as incidence rate ratios (IRR) and their 95% confidence intervals.

Univariable models were fitted with each pre-specified covariate as the independent variable and the primary and secondary outcomes as the dependent variables. Partially adjusted models were then fitted using a stepwise addition of covariates to explore the impact of different potential confounders. The stepwise addition of covariates included the demographic characteristics, followed by comorbidities, and medications. Fully adjusted models were then fitted with all pre-specified covariates. Age was only included as a covariate for adults aged 18 years and above. Smoking status was not included as a covariate for children aged 5 to 11 years due to the large amount of missing data. General practice was added as a random effect due to the variability in asthma treatment between general practices, such as differences in prescribing.

The population attributable fraction (PAF) was calculated to estimate the proportional reduction in asthma-related hospital admissions that would occur if exposure to the selected risk factors were eliminated from all people with asthma in a hypothetical scenario.^23^ The PAF for each selected modifiable risk factor was calculated in Stata using the ‘punaf’ command after running a fully adjusted negative binomial regression model including the selected risk factor. The PAF was only calculated for modifiable risk factors or treatable comorbidities that had statistically significant IRRs above 1.

### Study size

All eligible patients with asthma aged five years and above in the CPRD Aurum database were included. The sample size was determined by the available data.

### Missing data

Missing data were addressed by creating a separate ‘missing’ category for each binary or categorical variable, including ethnicity, IMD quintile, BMI and smoking status. This method was chosen in order to retain patient data and avoid introducing any biases from omitting patients with missing data from the analyses. The absence of clinical codes for diagnoses or drug treatment codes was taken to indicate the absence of the condition or drug.

### Data access and cleaning methods

Data were extracted using the Data Extraction for Epidemiological Research (DExtER)^24^ tool and the analyses were undertaken using Stata SE V.16 and RStudio. The dataset was inspected and cleaned to identify and remove any missing data, implausible values, and duplicate data. Summary statistics were generated for all covariates to identify implausible values. Histograms were generated for continuous variables and frequencies and proportions were generated for categorical variables. Duplicate data were removed from the dataset and implausible values were classified as ‘missing’ data. The HES APC linked dataset was merged with the CPRD Aurum master dataset. Patients without HES APC data linkage, or those not eligible for inclusion prior to the study start date were identified and removed from the dataset.

### Patient and Public Involvement

Patients and members of the public were not involved in the design or conduct of the study.

## Results

### Baseline characteristics

A total of 1,385,326 patients were included in the study (figure 1). This comprised 90,989 children, 114,927 adolescents, and 1,179,410 adults. The mean follow-up was 2.7 years.

**Figure 1.**
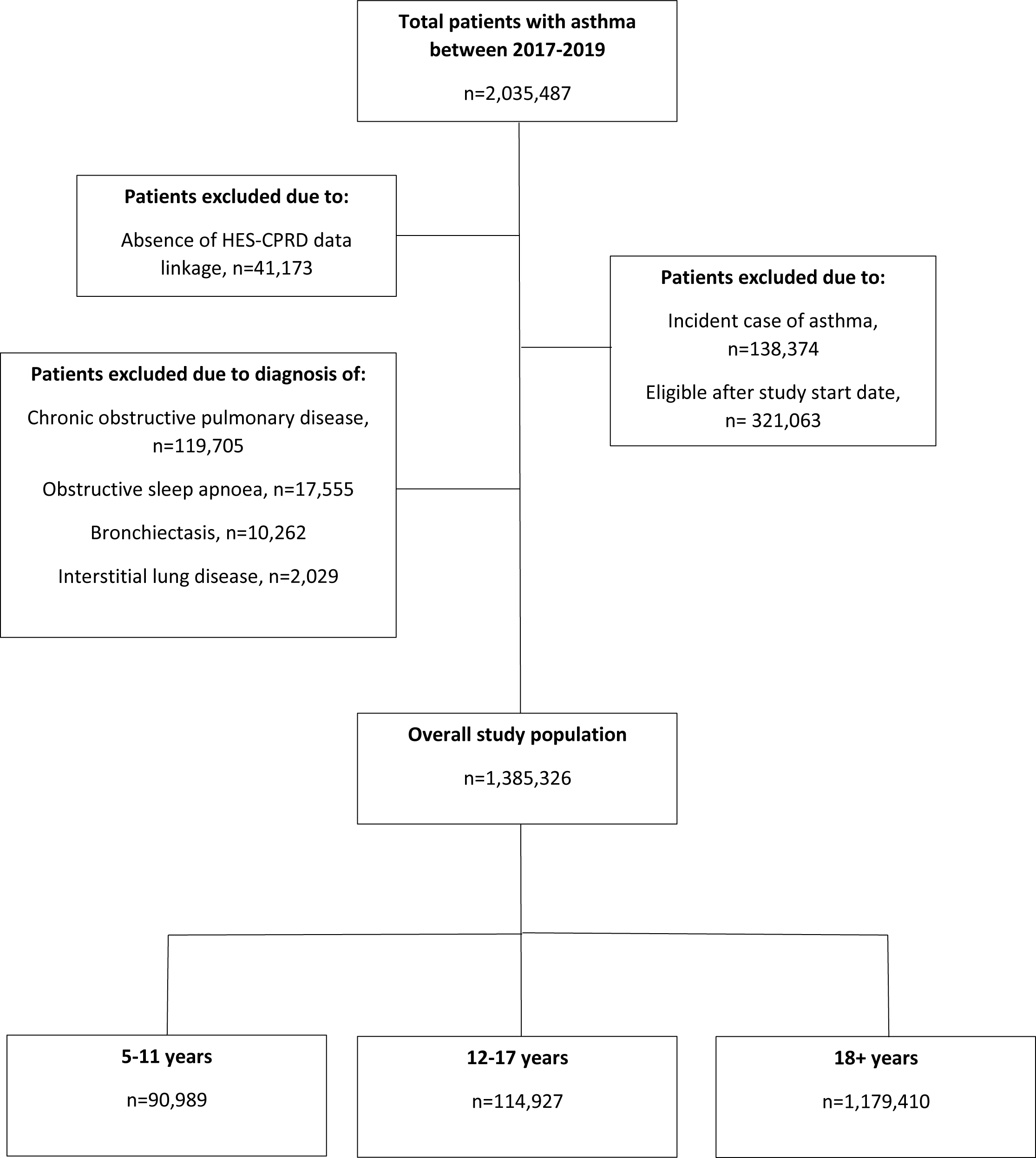
Flow diagram depicting study sampling and cohorts

Baseline characteristics, stratified by age group, are summarised in table 1. Among children, the median age was 9 years and 60% were male. 61% were of white ethnicity, 11% Asian and 5% black. Ethnicity data were missing for 18%. The proportion with normal weight was 35%, while 11% were either overweight or obese and BMI data were missing in 48%. The most common comorbidities were atopic eczema (45%), allergies (20%), allergic rhinitis (16%), and gastro-oesophageal reflux disease (6%). Most patients in this age group had been prescribed a SABA (65%), with 6% having received more than six prescriptions in the previous year. Only 54% were prescribed inhaled corticosteroids and 12% had been prescribed oral corticosteroids (4% were prescribed two or more) in the previous year. LTRAs were prescribed in 10% of cases, and only 37% had received an influenza vaccine in the previous year.

Among adolescents, the median age was 16 years and 59% were male. 49% were of white ethnicity, 8% Asian, 4% black and ethnicity data was missing in 35%. The proportion with normal weight was 33%, and 14% were overweight or obese and the BMI data was missing in 39%. Ten percent had a record indicating they were a current smoker and 10% a former smoker, with 26% having missing smoking data. Like children, the most common comorbidities were atopic eczema (44%), allergic rhinitis (26%), and allergies (26%), although the prevalence of anxiety and depression was higher (5% with anxiety and 3% with depression). 45% had been prescribed a SABA inhaler in the previous year and 5% had received over six such prescriptions during that period. 34% had been prescribed an inhaled corticosteroid, 4% a LTRA, and 6% at least one oral corticosteroid (2% were prescribed two or more) in the previous year, while 19% had received an influenza vaccine.

**Table 1.** Baseline characteristics.

| Characteristic | Children<br>(n=90,989) | Adolescents<br>(n=114,927) | Adults<br>(n=1,179,410) |
| --- | --- | --- | --- |
| Age (years), median (IQR) | 9.1 (7.4-10.6) | 15.5 (13.5-16.5) | 41.5 (29.5-56.5) |
| Age categories (years), n (%) |  |  |  |
| 18-24 | - | - | 175,957 (14.9) |
| 25-39 | - | - | 375,254 (31.8) |
| 40-59 | - | - | 392,801 (33.3) |
| 60-79 | - | - | 192,344 (16.3) |
| ≥80 | - | - | 43,054 (3.7) |
| Sex, n (%) |  |  |  |
| Male | 54,559 (60.0) | 67,825 (59.0) | 555,856 (47.1) |
| Female | 36,430 (40.0) | 47,102 (41.0) | 623,554 (52.9) |
| Ethnicity |  |  |  |
| White | 55,659 (61.2) | 56,275 (49.0) | 780,056 (66.1) |
| Black | 4,252 (4.7) | 4,527 (3.9) | 29,490 (2.5) |
| Mixed | 3,386 (3.7) | 2,905 (2.5) | 17,803 (1.5) |
| Asian | 9,834 (10.8) | 9,392 (8.2) | 68,409 (5.8) |
| Other | 1,375 (1.5) | 1,260 (1.1) | 11,304 (1.0) |
| Missing | 16,483 (18.1) | 40,568 (35.3) | 272,348 (23.1) |
| IIMD score, n (%) |  |  |  |
| 1 – Least deprived | 17,312 (19.0) | 22,938 (20.0) | 253,814 (21.5) |
| 2 | 15,788 (17.4) | 20,923 (18.2) | 239,399 (20.3) |
| 3 | 15,681 (17.2) | 20,377 (17.7) | 226,015 (19.2) |
| 4 | 18,686 (20.5) | 23,186 (20.2) | 233,339 (19.8) |
| 5 – Most deprived | 23,470 (25.8) | 27,404 (23.8) | 225,673 (19.1) |
| Missing | 52 (0.1) | 99 (0.1) | 1,170 (0.1) |
| BMI, n (%) |  |  |  |
| Underweight | 5,454 (6.0) | 15,859 (13.8) | 34,438 (2.9) |
| Normal weight | 31,496 (34.6) | 37,788 (32.9) | 379,089 (32.1) |
| Overweight | 7,151 (7.9) | 10,889 (9.5) | 334,266 (28.3) |
| Obese | 3,216 (3.5) | 5,604 (4.9) | 297,323 (25.2) |
| Missing | 43,672 (48.0) | 44,787 (39.0) | 134,294 (11.4) |
| Smoking status, n (%) |  |  |  |
| Current smoker | - | 11,749 (10.2) | 324,947 (27.6) |
| Former smoker | - | 11,970 (10.4) | 439,116 (37.2) |
| Never smoked | - | 60,832 (52.9) | 397,038 (33.7) |
| Missing |  | 30,376 (26.4) | 18,309 (1.6) |
| Comorbidities, n (%) |  |  |  |
| Allergies | 18,457 (20.3) | 29,658 (25.8) | 317,054 (26.9) |
| Atopic eczema | 40,808 (44.9) | 51,067 (44.4) | 317,822 (27.0) |
| Allergic rhinitis | 14,963 (16.4) | 29,492 (25.7) | 320,075 (27.1) |
| Gastro-oesophageal reflux disease (GORD) | 5,235 (5.8) | 3,503 (3.1) | 119,111 (10.1) |
| Chronic rhinosinusitis | 66 (0.1) | 243 (0.2) | 27,039 (2.3) |
| Anxiety | 946 (1.0) | 5,552 (4.8) | 260,258 (22.1) |
| Depression | 917 (1.0) | 2,949 (2.6) | 357,277 (30.3) |

| Medication use, n (%) |  |  |  |
| --- | --- | --- | --- |
| SABA | 59,321 (65.2) | 52,212 (45.4) | 464,123 (39.4) |
| SABA prescriptions within last year |  |  |  |
| 0 | 31,668 (34.8) | 62,715 (54.6) | 715,287 (60.7) |
| 1-3 | 42,416 (46.6) | 37,236 (32.4) | 288,395 (24.5) |
| 4-6 | 11,390 (12.5) | 8,822 (7.7) | 90,047 (7.6) |
| ≥7 | 5,515 (6.1) | 6,154 (5.4) | 85,681 (7.3) |
| OCS | 10,476 (11.5) | 6,333 (5.5) | 114,033 (9.7) |
| ICS | 48,865 (53.7) | 39,549 (34.4) | 445,349 (37.8) |
| LABA | 938 (1.0) | 1,245 (1.1) | 59,643 (5.1) |
| LAMA | 0 (0) | 6 (0.01) | 7,628 (0.7) |
| LTRA | 9,404 (10.3) | 4,920 (4.3) | 33,247 (2.8) |
| Influenza vaccine | 33,464 (36.8) | 22,256 (19.4) | 389,583 (33.0) |

BMI=Body mass index, ICS=inhaled corticosteroid, IQR=Interquartile range, LABA= Long-acting beta-2 agonists, LAMA=long-acting muscarinic antagonists, LTRA= Leukotriene receptor antagonists, OCS=oral corticosteroid, SABA=short acting beta-2 agonist

In the adult group, the median age was 42 years and 47% were male. 66% were of white ethnicity, followed by Asian (6%) and black (3%) and ethnicity data were missing for 23%. A significantly higher proportion of adults had BMI data compared to children and adolescents, with only 11% having missing data, and 54% being either overweight or obese. 28% were current smokers and 37% former smokers. The most common comorbidities were allergic rhinitis (27%), allergies (27%), atopic eczema (27%), depression (30%), and anxiety (22%). 39% had been prescribed a SABA inhaler in the previous year, with 7% having been prescribed more than six such inhalers during that period. 38% had been prescribed an inhaled corticosteroid, 10% an oral corticosteroid (4% were prescribed two or more), 6% had been prescribed either a LABA or LAMA, 3% a LTRA, and only 33% had received an influenza vaccine.

### Incidence rates of primary and secondary outcomes

During the study period, 2,898 of 90,989 (3.2%) children, 1,422 of 114,927 (1.2%) adolescents, and 13,186 of 1,179,410 (1.1%) adults experienced at least one incident asthma-related hospital admission during the three-year follow-up period. The distribution of the number of hospital admissions experienced by all patients with at least one hospital admission per year is shown in supplementary figure 1. Asthma-related hospital admission rates were highest in children (20.1 per 1000 person-years, 95% CI 19.5 to 20.6) compared to adolescents (10.1, 9.74 to 10.4) and adults (6.89, 6.79 to 6.98) (table 2). Similarly, ICU admissions were higher in children (0.54 per 1000 person-years, 95% CI 0.45 to 0.64) compared to adolescents (0.29, 0.23 to 0.35) and adults (0.15, 0.14 to 0.16) age categories (table 2).

**Table 2.** Incidence rates of primary and secondary outcomes. CI=confidence interval, ICU=intensive care unit

| Outcome | 5-11 years | 12-17 years | ≥18 years |
| --- | --- | --- | --- |
| **Hospital admissions** |  |  |  |
| Incident asthma hospital admissions | 5,105 | 3,228 | 21,925 |
| Person years | 254,526 | 320,086 | 3,184,183 |
| Incidence rate (per 1,000 person years) | 20.06 (95% CI 19.51-20.61) | 10.08 (95% CI 9.74-10.44) | 6.89 (95% CI 6.79-6.98) |
| **ICU admissions** |  |  |  |
| Incident ICU admissions | 137 | 92 | 474 |
| Person years | 254,526 | 320,086 | 3,184,183 |
| Incidence rate (per 1,000 person years) | 0.54 (95% CI 0.45-0.64) | 0.29 (95% CI 0.23-0.35) | 0.15 (95% CI 0.14-0.16) |

### Risk factors for hospital admissions

#### Children

Among children, males and females had a similar risk of asthma-related hospital admissions (IRR 0.95, 95% CI 0.87 to 1.04: figure 2 and supplementary table 2a). All ethnic minority groups including black (1.91,1.58 to 2.30), mixed (1.61,1.31 to 1.98), Asian (1.29, 1.11 to 1.49) and other (1.46, 1.05 to 2.02) ethnic groups were at an increased risk of hospital admissions compared to those from white ethnicity. Increasing socioeconomic deprivation was associated with an increased risk of hospital admissions, with those in the most deprived quintile having a 51% higher incidence rate than those in the least deprived quintile (1.51, 1.30 to 1.75).

**Figure 2.**
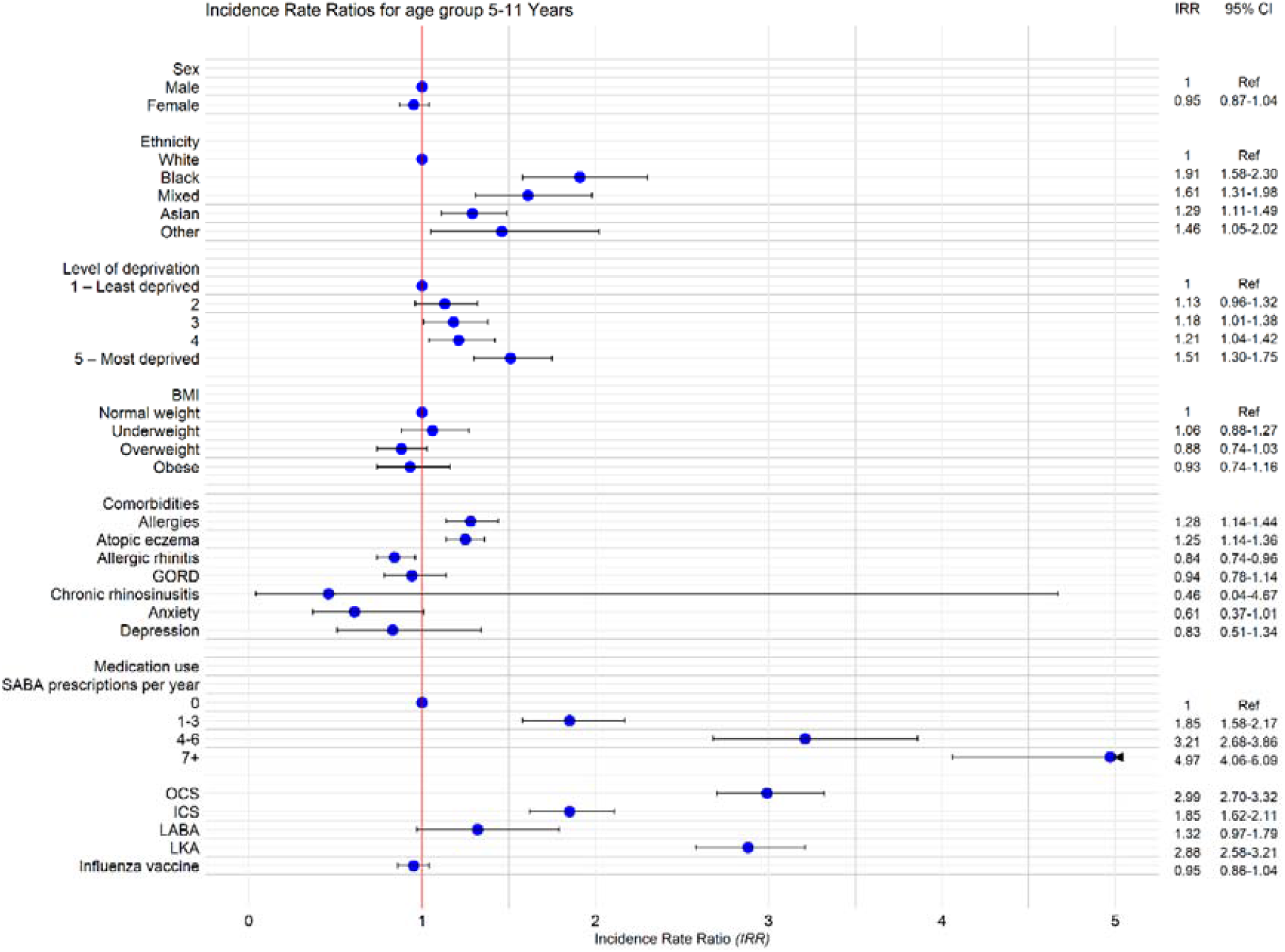
Adjusted Incidence Rate Ratios (IRR) for hospital admissions among children aged 5-11 years.

Overweight and obesity was not associated with an increased risk of hospital admissions compared to those of normal BMI. However, the presence of allergies (IRR 1.28, 95% CI 1.14 to 1.44) and atopic eczema (1.25, 1.14 to 1.36) were associated with an increased risk. Approximately 18% of asthma-related hospital admissions among children were attributable to atopic diseases including atopic eczema (PAF 11.4%, 95% CI 6.8 to 15.8) and allergies (6.8%, 95% CI 3.6 to 9.9) (see supplementary table 3). By contrast, allergic rhinitis was associated with a 16% decreased risk (0.84, 0.74 to 0.96).

Increasing number of SABA prescriptions was the factor most associated with an increased risk of hospital admissions, with those who had received more than six prescriptions in the previous year having a five-fold increased risk compared to those who had not received any (IRR 4.97, 95% CI 4.06 to 6.09). Previous prescriptions of other asthma medications were also associated with an increased risk of hospital admission. Those who had received oral corticosteroids or LTRAs were at almost three times the risk of hospital admissions compared to those who had not been prescribed them (IRR 2.99, 95% CI 2.70 to 3.32 and 2.88, 2.58 to 3.21, respectively).

#### Adolescents

Among adolescents, females had a greater risk of asthma-related hospital admissions than males (IRR 1.83, 95% CI 1.61 to 2.08: figure 3 and supplementary table 2b). All ethnic minority groups had an increased risk of hospital admissions compared to those of white ethnicity, including black (1.74, 1.31 to 2.32), mixed (1.74, 1.24 to 2.44), Asian (1.42, 1.14 to 1.78), and other (2.01, 1.21 to 3.34) ethnic groups. Like the younger age group, increasing socioeconomic deprivation was associated with an increased risk of hospital admissions among adolescents, with those in the most deprived quintile having a 52% increase in risk compared to the those from the least deprived quintile (1.52, 1.22 to 1.89).

**Figure 3.**
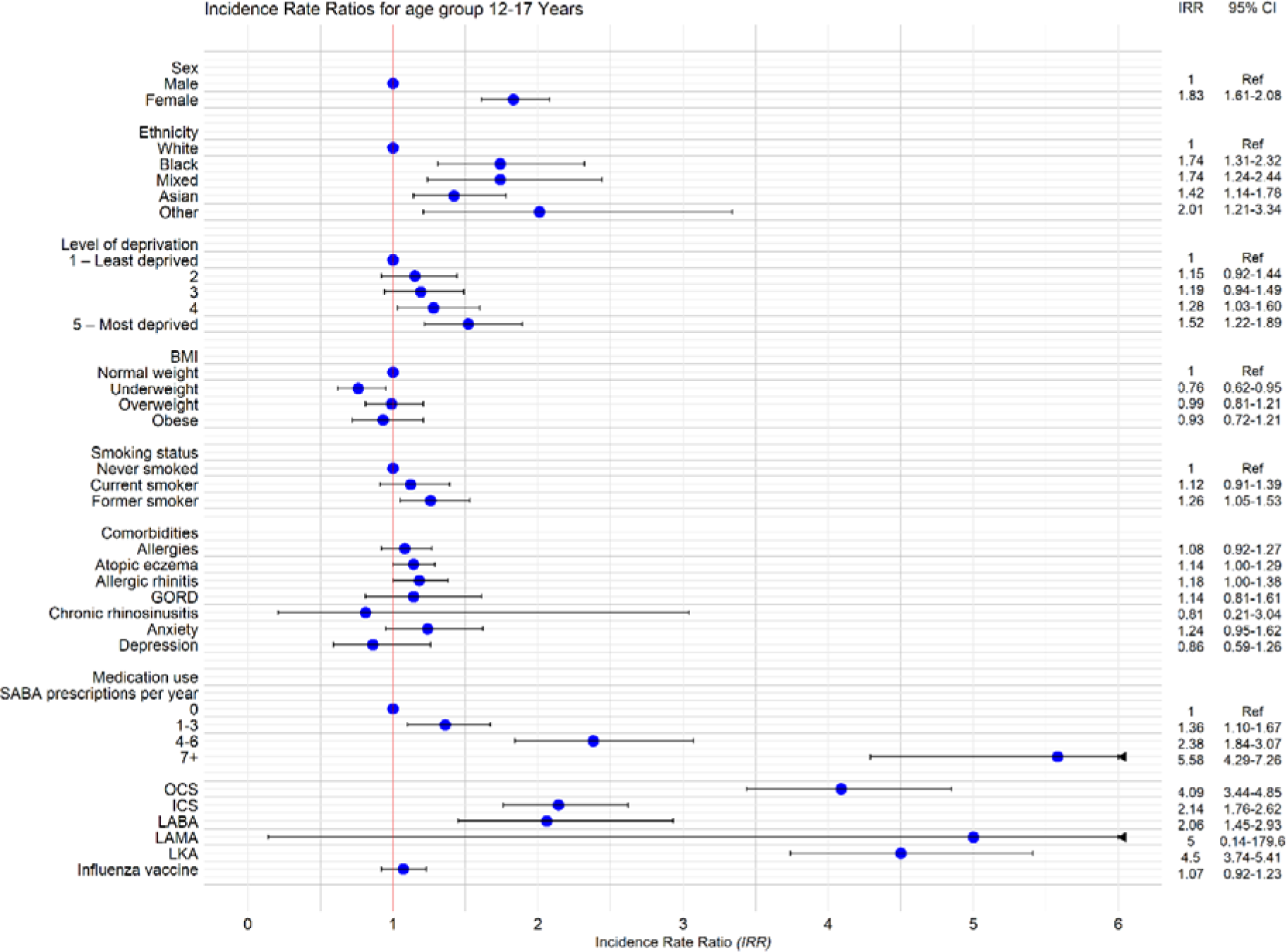
Adjusted Incidence Rate Ratios (IRR) for hospital admissions among adolescents aged 12-17 years. GORD; Gastro Oesophageal Reflux Disease, SABA; Short-Acting Beta-Agonist, OCS; Oral Corticosteroids, ICS; Inhaled Corticosteroids, LABA; Long-Acting Beta-Agonist, LAMA; Long-Acting Muscarinic Antagonist, LKA; Leukotriene Receptor Antagonist

High BMI was not a risk factor for hospital admissions, with overweight and obesity both having a similar risk to normal BMI, while being underweight was associated with a 24% lower risk than those of normal BMI (IRR 0.76, 95% CI 0.62 to 0.95). Former smoking was associated with a 26% increased risk of hospital admissions, compared to never smokers (1.26, 1.05 to 1.53). Approximately 7% of asthma-related hospital admissions were attributable to being a current/former smoker among adolescents (PAF 6.8%, 95% CI 0.9-12.3) (see supplementary table 3).

Allergic rhinitis was the only comorbidity that showed a statistically significant association with the risk of hospital admissions with an 18% increased risk (IRR 1.18, 95% CI 1.00 to 1.38). However, the associated PAF was not statistically significant.

Adolescents who had received prescriptions of several asthma-related medications were also at an increased risk of hospital admissions. This was particularly evident for those who had received an increasing number of SABA inhaler prescriptions, with those in receipt of more than six prescriptions in the previous year being at more than five times increased risk of hospital admissions than those who had not received any such prescriptions (IRR 5.58, 95% CI 4.29 to 7.26). Those who had been prescribed oral and inhaled corticosteroids (4.09, 3.44 to 4.85 and 2.14, 1.76 to 2.62, respectively), LABAs (2.06, 1.45 to 2.93), and LTRAs (4.50, 3.74 to 5.41) were also at an increased risk of hospital admissions.

#### Adults

Among adults, increasing age was associated with a decreasing risk of asthma-related hospital admissions compared to those aged 18 to 24 years, up until the age of 80 years. For example, those aged 60-79 years had a 42% decreased risk (IRR 0.58, 95% CI 0.53 to 0.62) compared to those aged 18 to 24 years, while those aged 80 years and older had a similar risk (1.06, 0.96 to 1.18) (see figure 4 and supplementary table 2c). Females were again at increased risk compared to males (1.61, 1.54 to 1.68). Compared to white adults, those of black (1.46, 1.30 to 1.64), mixed (1.25, 1.07 to 1.45) and Asian (1.60, 1.48 to 1.74) ethnic groups were at higher risk than those from white ethnic groups. Like the younger age groups, the risk of hospital admissions increased with levels of socioeconomic deprivation, with those in the most deprived quintile having a 43% higher risk of hospitalisation compared to those in the least deprived quintile (1.43, 1.33-1.54).

**Figure 4.**
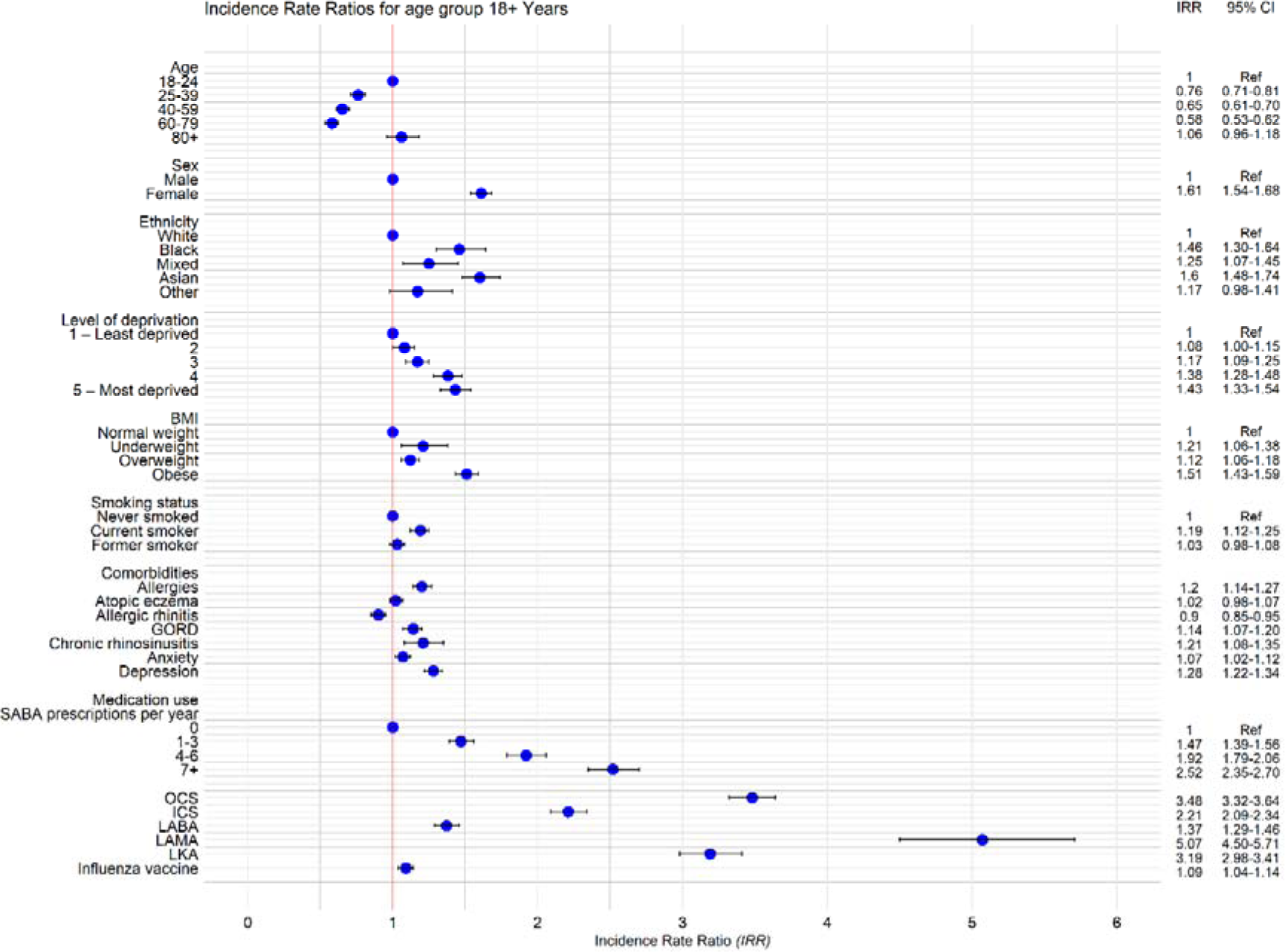
Adjusted Incidence Rate Ratios (IRR) for hospital admissions among adults aged 18+ years. GORD; Gastro Oesophageal Reflux Disease, SABA; Short-Acting Beta-Agonist, OCS; Oral Corticosteroids, ICS; Inhaled Corticosteroids, LABA; Long-Acting Beta-Agonist, LAMA; Long-Acting Muscarinic Antagonist LKA; Leukotriene Receptor Antagonist

Being underweight (IRR 1.21, 95% CI 1.06 to 1.38), overweight (1.12, 1.06 to 1.18) and obese (1.51, 1.43 to 1.59) were all associated with an increased risk of asthma-related hospital admissions compared to those of normal BMI. Similarly, current, but not former smokers were at increased risk of hospital admission compared to never smokers (1.19, 1.12 to 1.25). Several comorbidities were associated with an increased risk of hospital admissions, including allergies (IRR 1.20, 95% CI 1.14 to 1.27), GORD (1.14, 1.07 to 1.20), chronic rhinosinusitis (1.21, 1.08 to 1.35), anxiety (1.07, 1.02 to 1.12) and depression (1.28, 1.22 to 1.34). Allergic rhinitis by contrast was associated with a 10% decreased risk of hospital admissions (0.90, 0.85 to 0.95).

Medication prescriptions in the previous year were also associated with an increased risk of asthma-related hospitalisation. Those prescribed LAMAs had the highest increased risk of hospitalisation, with a five-fold increase in risk compared to those not prescribed LAMAs (IRR 5.07, 95% CI 4.50 to 5.71), although this represented a very small proportion of the cohort (0.7%). This was followed by prescription of oral corticosteroids, which was associated with a more than three-fold increased risk of hospital admissions (3.48, 3.32-3.64). Six or more SABA inhaler prescriptions was associated with more than a two-fold higher risk of hospital admission (2.52, 2.35-2.70) compared to having no SABA prescriptions. Prescription of inhaled corticosteroids, LABAs, LTRAs, and influenza vaccine were also all associated with an increased risk of hospital admission.

The highest proportion of asthma-related hospital admissions among adults was attributable to obesity (PAF 23.3%, 95% CI 20.5 to 26.1), followed by depression (11.1%, 9.1 to 13.1), allergies (6.2%, 4.4 to 8.0), smoking (4.3%, 3.0 to 5.7), GORD (2.3%, 1.2 to 3.4), anxiety (2.0%, 0.5 to 3.6) and chronic rhinosinusitis (0.8%, 0.3 to 1.3) (see supplementary table 3).

### Risk factors for ICU admissions

#### Children

Among children, those from black (IRR 4.07, 95% CI 2.35-7.05) and mixed (2.49, 1.23-5.05) ethnic groups were at a higher risk of asthma-related ICU admissions compared to those from white ethnic groups (see supplementary table 4a). High level of socioeconomic deprivation was also associated with an increased risk, with those in the most deprived quintile having a 98% increased risk (IRR 1.98, 95% CI 1.03 to 3.79) compared to those in the least deprived quintile.

Increasing number of SABA prescriptions in the previous year was strongly associated with the risk of ICU admissions, with those who had received more than six prescriptions having an approximately seven-fold increase in risk compared to those who had not received any (IRR 6.97, 95% CI 2.91 to 16.68). This was followed by prescriptions of LTRAs, which was associated with just under a three-fold increased risk (2.86, 1.93 to 4.24), while prescriptions of oral corticosteroids were associated with an approximately two-fold increase in risk (1.98, 1.34 to 2.92).

#### Adolescents

Adolescent females were at increased risk of asthma-related ICU admissions compared to males (IRR 1.54, 95% CI 1.00 to 2.36) (see supplementary table 4b). Black ethnicity was associated with more than a three-fold higher risk of ICU admissions compared to those from white ethnic groups (3.51, 1.62 to 7.59), while the differences observed in other ethnic groups were not statistically significant. The risk of asthma-related ICU admissions was increased by two-fold in the most derived socioeconomic quintile compared to those from the least deprived quintile (2.09, 0.88 to 4.98); however, the difference observed was not statistically significant.

Greater than six SABA prescriptions in the previous year was significantly associated with an eight-fold increased risk of ICU admissions in this age group compared to those without a prescription (IRR 8.44, 95% CI 2.49 to 28.56). Prescriptions of OCS (4.06, 2.56 to 6.45), ICS (3.95, 1.40 to 11.17) and LTRAs (2.77, 1.71 to 4.49) were all associated with an increased risk of asthma-related ICU admissions.

#### Adults

Among adults, increasing age was associated with a decreased risk of asthma-related ICU admissions, with those aged 80 years and older having an 81% decreased risk (IRR 0.19, 95% CI 0.10 to 0.38), compared to those aged 18 to 25 years (supplementary table 4c). Females were again at an increased risk of asthma-related ICU admissions compared to males (1.46, 1.17 to 1.81). Those from black, mixed, and Asian ethnic groups were at increased risk compared to those from white ethnic groups (IRR 2.69, 95% CI 1.75 to 4.13; 2.03, 1.10 to 3.74; and 1.46, 1.02 to 2.07, respectively). Depression was also associated with a 60% increased risk (1.60, 1.28 to 2.00).

Prescriptions of asthma medication in the previous year were also associated with an increased risk of asthma-related ICU admissions. There was a gradient of increasing risk of ICU admissions with increasing SABA prescriptions, with six or more in the previous year being associated with a four-fold higher risk (IRR 4.39, 95% CI 3.02 to 6.37) compared to having no SABA prescriptions. Similarly, OCS, ICS (3.33, 2.68 to 4.14; and 1.90, 1.36 to 2.67), LABAs (1.31, 1.01 to 1.70), LAMAs (4.28, 3.05 to 6.01), and LTRAs prescriptions (3.17, 2.46 to 4.07) were associated with an increased risk of asthma-related ICU admissions (see supplementary table 4c).

## Discussion

### Main findings

Our analysis of over one million patients showed that there are significant associations between age, sex, ethnicity, socioeconomic deprivation, and comorbidities, and asthma-related hospital and ICU admissions. 3% of children and 1% of all adolescents and adults with asthma had an asthma-related hospital admission within a three-year period.

In all age cohorts, ethnic minority groups particularly those of black and mixed ethnicity were at significantly increased risk of asthma-related hospital and ICU admissions compared to those from white ethnic groups. Socioeconomic deprivation was also a risk factor for asthma-related hospital and ICU admissions, with people in the lowest socioeconomic groups bearing a disproportionate burden of these admissions.

Children with allergies and atopic eczema were at increased risk of hospital admissions. Effective treatment of allergies and atopic conditions could potentially reduce asthma-related hospital admissions. Among adolescents, females were at higher risk than males for asthma-related hospital and ICU admissions. Smoking was also an important risk factor in this age group, and we estimated that 7% of asthma-related hospital admissions could be avoided by eliminating smoking in in this age group.

Among adults, women and people of younger age were at increased risk of asthma-related hospital and ICU admissions. Being underweight, overweight and obese were all associated with increases in risk of hospital admissions. Several comorbidities were also associated with an increased risk of hospital admissions, including allergies, gastro-oesophageal reflux disease, chronic rhinosinusitis, anxiety, and depression. Depression was particularly associated with an increased risk of ICU admissions in this age group. A significant proportion of all asthma-related hospital admissions in adults were attributable to modifiable risk factors, including obesity (approximately one fifth of all admissions) and depression (approximately one tenth), followed by allergies, smoking, gastro-oesophageal reflux disease, anxiety, and chronic rhinosinusitis.

Allergic rhinitis was associated with an increased risk of asthma-related hospital admissions among adolescents, highlighting the importance of addressing this comorbidity. Counterintuitively, in children and adults it was associated with a reduced risk in the primary outcome in the fully adjusted negative binomial models. However, the risk was increased in the univariable analyses and the reduction in the incidence rate ratios may be attributable to adjustment for other atopic diseases.

In all three age cohorts, previous prescriptions of asthma medicines were significantly associated with an increased risk of hospital and ICU admissions. Increasing SABA prescriptions in the previous year was associated with a gradient of increasing risk in hospital and ICU admissions in a dose dependent manner. More than six such prescriptions were especially associated with increased risk, particularly in children and adolescents. Prescriptions of OCS, ICS, LABAs, LAMAs, and LTRAs, were associated with an increased risk of hospital and ICU admissions. These associations are likely to reflect increasing disease severity (since patients with more severe disease receive a wider range of asthma treatments), poor symptom control (a higher number of SABA and oral corticosteroid prescriptions are likely to represent uncontrolled asthma), and potentially poor medication compliance (e.g., high use of SABAs may be indicate inadequate compliance with preventer medication).

### Relationship to other studies

The relationship between age and asthma related morbidity is inconsistent between the current and previous studies. In contrast to our findings, some previous studies have demonstrated an increasing risk of hospital admissions, readmissions, and severe exacerbations with age.^25 26^ This differs from our findings which show that the highest incidence rates for hospital and ICU admissions are in children and adolescents. Among adults, the risk of hospitalisation decreased with increasing age up until 80 years and older, when the risk was similar to those aged 18 to 24 years. Previous studies have shown that older adults are more likely to have more severe asthma,^27^ poorer asthma control, ^28^ poorer response to asthma medications such as bronchodilators,^29^ and reduced lung function^30^ compared to younger individuals with asthma. On unadjusted analyses we similarly found that the risk of asthma-related hospital admissions increased with increasing age (supplementary table 2c). However, once adjusted for demographic and behavioural risk factors (BMI and smoking), the relationship between age and hospital admissions mostly inverted, suggesting that the positive association was largely driven by confounders.

As observed in our study, previous studies have suggested that female sex is an important risk factor for asthma-related hospital admissions. Two studies that included both children and adults with asthma found an association between female sex and a higher risk of hospital admissions.^31 32^ Another study based in Canada that used data from 86,863 young people under 20 years of age between 1994 and 1997 found that females aged 15-19 years had an increased risk of asthma-related readmissions compared to males.^33^ Similarly, a more recent study that used data from 15,691 patients registered in national Swedish health registries between 2006 and 2015 found that females had an increased risk of being re-hospitalised across all age groups.^34^ The reasons for these sex differences is unclear but there is some evidence that ovarian hormones may increase and testosterone may decrease airway inflammation in asthma, although further research is needed to clarify these pathways.^35^ This is supported by the finding that this sex difference in risk was not seen in girls aged 5-11 years in our study. However, one study found that girls with asthma under the age of 12 years had a statistically significant increase in the risk of readmission to hospital compared to males, which contrasts with our findings.^36^

Ethnic disparities in asthma-related hospital admissions and re-admissions have been found in several previous studies, particularly in patients from black ethnic minority groups who appear to have the highest risk among children.^37 38 39^ Differences in socioeconomic status may partially explain these ethnic disparities. Beck et al (2014) found that social hardships and socioeconomic status explained approximately 40-50% of the racial disparity observed for paediatric asthma-related hospital admissions in one study in the United States.^39^ A lower socioeconomic status may translate into less favourable socioeconomic conditions such as reduced access to healthcare and poor living conditions, such as proximity to higher levels of air pollution. In addition, higher BMI, airway inflammatory responses to early life exposures, and genetic predisposition have been implicated in increasing the risk of asthma exacerbations in ethnic minority children.^40^

Several studies have previously demonstrated that increased socioeconomic deprivation is a risk factor for asthma-related hospital admissions and re-admissions in children and adults.^41 42 43^ The risk of hospital admissions has previously been shown to be between one and two times higher in those who are most deprived compared to those who are least. This is consistent with the findings from our study and may be explained by the many adverse factors that are associated with low socioeconomic status such as poor housing conditions.^44^ This effect may be particularly marked in urban areas.^42^

Studies have investigated the association between exposure to tobacco smoke and risk of hospitalisation, readmission, and severe asthma exacerbations.^45 46 47^ Two studies found that detectable serum or salivary cotinine was significantly associated with a 59% and 135% increase in asthma-related hospital readmissions, respectively,^45^ and a 40% increase in severe asthma exacerbations (defined as hospitalisation, emergency department visit or prescription of an oral corticosteroid).^47^

Several studies have found that obesity is associated with an up to four-fold increased risk of asthma-related hospitalisations and readmissions in children and adults,^48 49 50^ whereas in our study, we only found this association in adults. The lack of association between obesity and asthma-related hospitalisations in younger people in our study could be partly attributable to the high level of missing BMI data for children and adolescents. However, a meta-analysis of observational studies similarly found that overweight and obesity is not associated with asthma-related hospitalisations in children and adolescents.^51^ The association between obesity and increased risk in hospital admissions in adults may be explained by the inflammatory effects of obesity and the impact on airway caliber and bronchial hyper-reactivity.^52 53 54^

Previous studies have demonstrated an association between affective disorders, such as anxiety and depression, and the risk of asthma-related emergency department (ED) visits and hospitalisation.^55 56 57^ One cross-sectional study of adults with asthma that captured data using the Asthma Quality of Life Questionnaire showed a significant association between depression and asthma-related hospital attendances.^57^ Another study used data from 568 participants who participated in baseline and follow-up surveys, which showed that depression increased the risk for ED visits by approximately two-fold.^56^ However, there is a scarcity of literature on the role of depressive disorders in children in the risk of asthma hospitalisations, which may be partly due to depressive symptoms being relatively undiagnosed or untreated in children.^58 59 60^

A recent systematic review and meta-analysis of observational studies showed that gastro-oesophageal reflux disease is a risk factor for severe asthma.^61^ The findings from the meta-analysis showed that the overall odds of severe asthma exacerbations were 27% higher in those with GORD. Furthermore, children with GORD were more at risk of severe asthma exacerbations than adults.^61^ This is in contrast to our study, which found an increased risk in asthma-related hospitalisations associated with GORD only in adults and not in children or adolescents. This may potentially be due to under recognition of GORD in children and adolescents, leading to exposure misclassification bias in our analysis and dilution of the associated effect size. However, it is also possible that GORD plays a more dominant role in asthma outcomes in adults, in which it is a more common comorbidity, and may be less significant in children. We would expect GORD to be associated with worse asthma control since acid reflux can induce cough and induce airway inflammation through pulmonary aspiration, and vagus-mediated bronchoconstriction may occur in response to reflux, worsening asthma symptoms.^62^ However, it remains unclear whether treating GORD has a significant impact on asthma exacerbations.

The presence of allergies or atopic illnesses were associated with asthma-related hospitalisations in all three age groups in our study. The role of allergens in triggering IgE-mediated inflammation, eosinophilia, and release of inflammatory cytokines, to promote airway inflammation and bronchial mucous hypersecretion in asthma, are well established.^63^ Both indoor and outdoor allergens are implicated in increased asthma related morbidity and mortality. However, evaluation of allergy is usually performed in patients with severe asthma. Our findings suggest that more effective management of allergies may result in reducing asthma related hospitalisation in children and adults. ^64 65 66^ However, in practice it can often be challenging to reduce exposure to allergens.

There are number of studies in the literature which support our findings that previous use of SABAs and oral corticosteroids are associated with an increased risk in hospital admissions.^67 68 37^ A study by Butz et al. showed that SABA use was associated with a two-fold increase in asthma-related hospitalisations,^69^ while Hull et al. and Stanford et al. both showed that the use of three or more SABA inhalers per year was associated with up to a three-fold increase in risk of asthma-related exacerbations.^37 70^ It is well documented in the literature that frequent SABA use is an indicator of poor asthma control, which is likely to explain the increased hospital admission risk with increasing SABA prescriptions observed in the current study.^67 71 72^ Medication burden in general is a marker of poor asthma control, potentially poor medication compliance, and greater disease severity. A longitudinal study which followed over 1000 New Zealanders from birth to 26 years of age similarly found that asthma-related hospital admissions were overall more likely in patients who had been treated with a SABA, inhaled corticosteroid, or any other asthma medication. ^74^

### Strengths and limitations

The study used linked primary and secondary care data from over a million patients with diagnosed asthma, making this one of the largest analyses of asthma-related hospital and ICU admissions. The large sample size provided adequate power to investigate several important risk factors, and stratify analyses by age subgroups, including children, adolescents and adults. We evaluated a range of risk factors for asthma-related hospital and ICU admissions and estimated the population attributable fraction for key modifiable risk factors. This enabled us to identify the most important risk factors responsible for these outcomes at a population level.

There were also several limitations to the study. We defined asthma as a coded diagnosis of asthma using pre-specified SNOMED-CT terms that had been selected using a systematic process. However, asthma is commonly misdiagnosed and underdiagnosed in primary care.^75^ However, previous research has demonstrated that asthma can be accurately defined within electronic health record data.^76^ There may also be misclassification bias in the coding of asthma-related hospital admissions, particularly in adults, as exacerbations could be due to other comorbidities with overlapping symptoms, such as heart failure. However, we attempted to limit this by excluding patients with common chronic respiratory diseases such as COPD, which might otherwise present similarly to asthma during acute exacerbations.

The study was also limited by the lack of data on some key risk factors, such as BMI and smoking status in children and adolescents, as well as lacking data on environmental risk factors such as air quality. Although we had accurate data on prescriptions, we did not have any data on medication compliance. We also lacked data on key physiological measures such as peak expiratory flow rate, spirometry, airway eosinophilia, fractional exhaled nitric oxide (FeNO), and other physiological measures of disease severity, or patient reported outcome measures such as the Asthma Control Test.

We were primarily interested in assessing modifiable risk factors potentially driving future asthma-related hospital and ICU admissions. Although previous asthma-related hospitalisations are a risk factor for future exacerbations, we did not include this in our models since this would not add to our understanding of modifiable risk factors and would potentially lead to overadjustment in our regression models. Finally, although we were able to include outcome data on hospital and ICU admissions, we did not have access to data on emergency department attendances, which also constitutes an important measure of asthma control and secondary care use.

### Implications for practice, policy, and future research

Our study provides novel insights into age specific risk factors and population groups experiencing severe asthma exacerbations requiring hospital and ICU admission. We found significant ethnic and socioeconomic disparities in asthma outcomes, particularly with respect to children from black ethnic minority backgrounds. These disparities require urgent attention by health service commissioners and providers to promote improvements in asthma management in disadvantaged population groups. Work should also be done by public health services to highlight this inequality and tackle the social and environmental factors that may be contributing to poor asthma outcomes in socioeconomically deprived and ethnic minority communities. In addition, younger people and women are other demographic groups disproportionately affected by poor asthma control. This should also be considered when designing public health approaches to improving asthma outcomes and in the design of clinical services for asthma.

We have provided estimates of the proportion of asthma-related hospital admissions that could potentially be prevented by eliminating modifiable risks factors. Our findings suggest that treating allergies and atopic conditions should be considered an important component of asthma management and has the potential to significantly reduce asthma-related hospital admissions in all age groups. In adolescents and adults, smoking cessation also has the potential to significantly reduce hospital admissions, emphasizing the need to integrate smoking cessation services into asthma care. In adults, supporting weight management and treating comorbid depression, anxiety, and GORD, should also be considered an integral part of general asthma management as they are likely to significantly contribute to avoidable asthma-related hospital admissions.

We also show the importance of medication burden as a key measure of poor asthma control. In addition to monitoring SABA prescriptions, clinicians should also monitor the prescription frequency of other asthma medications including oral corticosteroids. High medication burden should be considered an important measure of disease severity and predictor of asthma-related hospital and ICU admissions. Measures of global asthma medication burden, incorporating the findings of this study, could be used by electronic prescribing systems to highlight high risk patients requiring regular review and support.

Research is needed to understand how to reduce demographic and social inequalities in asthma outcomes, including whole system public health approaches. Further research is also needed to evaluate the effectiveness and cost-effectiveness of integrating screening and treating the modifiable risk factors and comorbidities identified in this study. Research is also needed to better understand the relationship between obesity and asthma outcomes in children, and how weight management can be incorporated into asthma care in adults.

Future risk prediction modelling for asthma should consider the data on risk factors from this study. Beyond this, there is a need for impact trials on risk prediction models for asthma outcomes in primary care that include children, adolescents and adults. Finally, there is a need to develop global measures of asthma medication burden to support electronic prescribing systems to highlight high risk patients, and the utility of these measures on clinical care and outcomes should be evaluated.

## Conclusions

There remain significant sociodemographic inequalities in the rates of asthma-related hospital and ICU admissions. Key addressable risk factors are allergic and atopic conditions and smoking across all age groups. In adults, addressing obesity, depression, anxiety, and gastro-oesophageal reflux disease has the potential to significantly reduce asthma-related hospitalisations. Integrating the screening and treatment of these risk factors could improve asthma outcomes and reduce avoidable hospital admissions but this needs to be evaluated in randomised controlled trials. Finally, the overall asthma medication burden is an important measure of asthma severity and predictor of poor outcomes, which should form a key measure for asthma monitoring and support.

## Data Availability

Access to anonymised patient data from CPRD is subject to a data sharing agreement containing detailed terms and conditions of use following protocol approval from the MHRA Independent Scientific Advisory Committee. This study-specific analysable dataset is therefore not publicly available but can be requested from the corresponding author subject to research data governance approvals. Details about Independent Scientific Advisory Committee applications and data costs are available on the CPRD website (cprd.com).

## Figures

Figure 1 – Participant flow diagram

Figure 2 – Forest plot showing adjusted incidence rate ratios (IRR) for hospital admissions among children

Figure 3 – Forest plot showing adjusted incidence rate ratios (IRR) for hospital admissions among adolescents

Figure 4 - Forest plot showing adjusted incidence rate ratios (IRR) for hospital admissions among adults

