## Supplementary file for "Risk factors for asthma-related hospital and intensive care admissions in children, adolescents, and adults: a cohort study using primary and secondary care data"

**Supplementary Methods**

***Appendix 1***

**Poisson and Negative Binomial Regression for count data**

1. **Poisson Distribution**

The Poisson distribution is given by P(X=x) = $\frac{\lambda^{x}e^{-\lambda}}{x!}$ , X will have a Poisson distribution with parameter λ.

Where, X= number of counts (events) occurring in a fixed period of time, x= 0, 1, 2, 3… and the intensity parameter λ represents the expected number of occurrences in a fixed period of time (E(X)=λ) and it is also the variance of the counts (Var(X)=λ).

2. **Poisson Regression Model**

The goal is to model count data as a function of covariates. Given the Poisson distribution, we model the logarithm of its expected value (log (λ)) as a function of covariates. This creates the Poisson Regression Model as given below,

$$\log\left( E\left( Y | x \right) \right)=log(\lambda)=\alpha+ \beta^{T}x$$

3. **Negative Binomial Distribution**

The Negative binomial distribution is written as P(X=x) =$\left. \left( \begin{aligned} x-1 \\ r-1 \end{aligned} \right. \right){(1-p)}^{x-r}p^{r}$

Where, X = total number of trials, r = number of occurrences of success and p= probability of failure on each occurrence.

4. **Negative Binomial regression Model**

The negative binomial regression is similar to regular multiple regression except that the response variable is an observed count that follows the negative binomial distribution. The negative binomial model is also similar to the Poisson model, but incorporates an additional term to account for the excess variance. The negative binomial model can be used when variance is substantially higher than the mean.

Both Poisson and negative binomial regression give the same results in the absence of overdispersion. However negative binomial regression is more appropriate in the presence of overdispersion, and therefore we use negative binomial regression instead of Poisson regression in this study.

**Supplementary tables**

***Supplementary table 1***

**Asthma clinical code list**

| **DESCRIPTION** | **SNOMED-CT CODE** |
| --- | --- |
| Occupational asthma | 57607007 |
| Asthma | 195967001 |
| Brittle asthma | 225057002 |
| Intrinsic asthma | 266361008 |
| Asthma - currently active | 312453004 |
| Mild asthma | 370218001 |
| Allergic asthma | 389145006 |
| Asthma annual review | 394700004 |
| Asthma medication review | 394720003 |
| Extrinsic asthma - atopy | 389145006 |
| Occupational asthma | 955631000006100 |
| Acute non-infective exacerbation of asthma | 2010041000006100 |
| Asthma accident and emergency attendance since last visit | 708373002 |
| Emergency asthma admission since last appointment | 708358003 |
| Intrinsic asthma NOS | 266361008 |
| Late-onset asthma | 233679003 |
| Extrinsic asthma with asthma attack | 708093000 |
| Asthma attack NOS | 708038006 |
| Exercise induced asthma | 31387002 |
| Exercise induced asthma | 31387002 |
| Extrinsic (atopic) asthma | 389145006 |
| Acute exacerbation of asthma | 708038006 |
| Severe asthma attack | 708090002 |
| Status asthmaticus NOS | 734904007 |
| Emergency admission, asthma | 183478001 |
| Emergency admission, asthma | 183478001 |
| Life threatening acute exacerbation of non-allergic asthma | 1086711000000100 |
| Asthmatic bronchitis | 405944004 |
| Acute exacerbation of extrinsic asthma | 708093000 |
| Industrial asthma | 57607007 |
| EIA - Exercise-induced asthma | 31387002 |
| Emergency hospital admission for asthma | 183478001 |
| Acute exacerbation of asthma | 708038006 |
| Acute severe exacerbation of asthma | 708090002 |
| Life threatening acute exacerbation of extrinsic asthma | 1086701000000100 |
| Non-allergic asthma | 266361008 |
| Acute exacerbation of immunoglobulin E-mediated allergic asthma | 708093000 |
| IgE mediated allergic asthma | 424643009 |
| Immunoglobulin E-mediated allergic asthma | 424643009 |
| Exercise induced asthma | 31387002 |
| Asthmatic | 195967001 |
| Atopic asthma | 389145006 |
| Difficult asthma | 2009981000006100 |
| Occasional asthma exacerbations | 1821521000006100 |
| Moderate asthma | 370219009 |
| Occasional asthma | 370220003 |
| Hay fever with asthma | 233683003 |
| Mixed asthma | 195977004 |
| Extrinsic asthma NOS | 424643009 |
| Allergic asthma NEC | 389145006 |
| Severe asthma attack | 708038006 |
| IgE-mediated allergic asthma | 424643009 |
| IgE mediated asthma | 424643009 |
| Chronic asthma with fixed airflow obstruction | 866881000000101 |
| Severe asthma | 370221004 |
| Asthma follow-up | 394701000 |
| Chronic asthmatic bronchitis | 195949008 |
| Bronchial asthma | 195967001 |
| Asthma causing night waking | 170632009 |
| Asthma not disturbing sleep | 170635006 |
| Late-onset asthma | 233679003 |
| Asthma management plan given | 406162001 |
| Asthma unspecified | 195967001 |
| Asthma self-management plan agreed | 811921000000103 |
| Acute infective exacerbation of asthma | 2010031000006100 |
| Asthma confirmed | 401193004 |
| Asthma attack | 708038006 |
| Late onset asthma | 233679003 |
| Asthma severely restricts exercise | 170657004 |
| Intrinsic asthma with asthma attack | 708094006 |
| Asthma NOS | 195967001 |
| Intrinsic asthma with status asthmaticus | 1086711000000100 |
| Hay fever with asthma | 233683003 |

| **5-11 years**  ***Adjusted Incidence Rate Ratios (IRR) for hospital admissions among children aged 5-11 years***  **Supplementary table 2a** | | | | | | | | | | | | |
| --- | --- | --- | --- | --- | --- | --- | --- | --- | --- | --- | --- | --- |
|  | Univariable model | | | Multivariable models | | | | | | | | |
|  |  |  |  | Partially adjusted* | | | Partially adjusted** | | | Fully adjusted *** | | |
| Covariate | Unadjusted IRR | 95% CI | P-value | Adjusted IRR | 95% CI | P-value | Adjusted IRR | 95% CI | P-value | Adjusted IRR | 95% CI | P-value |
| Sex, n (%) |  |  |  |  |  |  |  |  |  |  |  |  |
| Male | 1 | - | - | 1 | - | - | 1 | - | - | 1 | - | - |
| Female | 0.92 | 0.84-1.01 | 0.073 | 0.89 | 0.81-0.98 | 0.018 | 0.90 | 0.82-0.99 | 0.032 | 0.95 | 0.87-1.04 | 0.263 |
| Ethnicity |  |  |  |  |  |  |  |  |  |  |  |  |
| White | 1 | - | - | 1 | - | - | 1 | - | - | 1 | - | - |
| Black | 2.09 | 1.71-2.55 | <0.001 | 1.90 | 1.54-2.35 | <0.001 | 1.73 | 1.40-2.13 | <0.001 | 1.91 | 1.58-2.30 | <0.001 |
| Mixed | 1.93 | 1.54-2.42 | <0.001 | 1.71 | 1.36-2.15 | <0.001 | 1.61 | 1.28-2.02 | <0.001 | 1.61 | 1.31-1.98 | <0.001 |
| Asian | 1.46 | 1.26-1.68 | <0.001 | 1.41 | 1.21-1.66 | <0.002 | 1.28 | 1.09-1.50 | 0.002 | 1.29 | 1.11-1.49 | 0.001 |
| Other | 1.34 | 0.93-1.92 | 0.117 | 1.36 | 0.95-1.96 | 0.093 | 1.35 | 0.94-1.94 | 0.100 | 1.46 | 1.05-2.02 | 0.023 |
| Missing | 0.96 | 0.85-1.09 | 0.524 | 1.03 | 0.90-1.18 | 0.624 | 1.02 | 0.90-1.17 | 0.725 | 1.01 | 0.89-1.15 | 0.836 |
| IMD score, n (%) |  |  |  |  |  |  |  |  |  |  |  |  |
| 1 – Least deprived | 1 | - | - | 1 | - | - | 1 | - | - | 1 | - | - |
| 2 | 1.44 | 1.23-1.69 | <0.001 | 1.24 | 1.05-1.46 | 0.013 | 1.23 | 1.04-1.46 | 0.013 | 1.13 | 0.96-1.32 | 0.141 |
| 3 | 1.35 | 1.15-1.59 | <0.001 | 1.24 | 1.05-1.47 | 0.012 | 1.25 | 1.05-1.47 | 0.010 | 1.18 | 1.01-1.38 | 0.042 |
| 4 | 1.58 | 1.35-1.83 | <0.001 | 1.33 | 1.13-1.57 | 0.001 | 1.37 | 1.17-1.62 | <0.001 | 1.21 | 1.04-1.42 | 0.015 |
| 5 – Most deprived | 2.19 | 1.90-2.53 | <0.001 | 1.75 | 1.49-2.05 | <0.001 | 1.79 | 1.53-2.10 | <0.001 | 1.51 | 1.30-1.75 | <0.001 |
| Missing | 1.61 | 0.24-10.57 | 0.621 | 1.59 | 0.25-10.33 | 0.627 | 1.98 | 0.32-12.47 | 0.466 | 2.96 | 0.58-15.17 | 0.194 |
| BMI, n (%) |  |  |  |  |  |  |  |  |  |  |  |  |
| Normal weight | 1 | - | - | 1 | - | - | 1 | - | - | 1 | - | - |
| Underweight | 1.00 | 0.82-1.22 | 0.991 | 0.96 | 0.79-1.17 | 0.699 | 0.99 | 0.81-1.20 | 0.907 | 1.06 | 0.88-1.27 | 0.558 |
| Overweight | 0.98 | 0.82-1.17 | 0.818 | 0.96 | 0.80-1.15 | 0.666 | 0.96 | 0.81-1.15 | 0.682 | 0.88 | 0.74-1.03 | 0.117 |
| Obese | 1.17 | 0.92-1.49 | 0.208 | 1.12 | 0.88-1.43 | 0.358 | 1.18 | 0.92-1.50 | 0.186 | 0.93 | 0.74-1.16 | 0.520 |
| Missing | 0.73 | 0.66-0.81 | <0.001 | 0.77 | 0.69-0.85 | <0.001 | 0.82 | 0.73-0.91 | <0.001 | 1.13 | 1.02-1.25 | 0.022 |
| Comorbidities, n (%) |  |  |  |  |  |  |  |  |  |  |  |  |
| Allergies | 1.77 | 1.59-1.97 | <0.001 | - | - | - | 1.55 | 1.36-1.75 | <0.001 | 1.28 | 1.14-1.44 | <0.001 |
| Atopic eczema | 1.68 | 1.53-1.84 | <0.001 | - | - | - | 1.56 | 1.42-1.71 | <0.001 | 1.25 | 1.14-1.36 | <0.001 |
| Allergic rhinitis | 1.35 | 1.20-1.52 | <0.001 | - | - | - | 0.97 | 0.84-1.11 | 0.649 | 0.84 | 0.74-0.96 | 0.008 |
| Gastro-oesophageal reflux disease (GORD) | 1.03 | 0.85-1.25 | 0.767 | - | - | - | 1.13 | 0.93-1.37 | 0.230 | 0.94 | 0.78-1.14 | 0.545 |
| Chronic rhinosinusitis | 0.27 | 0.02-2.93 | 0.282 | - | - | - | 0.33 | 0.03-3.56 | 0.361 | 0.46 | 0.04-4.67 | 0.509 |
| Anxiety | 0.68 | 0.42-1.10 | 0.115 | - | - | - | 0.59 | 0.36-0.98 | 0.043 | 0.61 | 0.37-1.01 | 0.054 |
| Depression | 0.85 | 0.53-1.37 | 0.512 | - | - | - | 0.94 | 0.57-1.57 | 0.821 | 0.83 | 0.51-1.34 | 0.447 |
| Medication use, n (%)† |  |  |  |  |  |  |  |  |  |  |  |  |
| Number of SABA prescriptions within last year, n (%) |  |  |  |  |  |  |  |  |  |  |  |  |
| 0 | 1 | - | - | - | - | - | - | - | - | 1 | - | - |
| 1-3 | 3.90 | 3.43-4.45 | <0.001 | - | - | - | - | - | - | 1.85 | 1.58-2.17 | <0.001 |
| 4-6 | 11.44 | 9.87-13.27 | <0.001 | - | - | - | - | - | - | 3.21 | 2.68-3.86 | <0.001 |
| 7+ | 25.57 | 21.61-30.24 | <0.001 | - | - | - | - | - | - | 4.97 | 4.06-6.09 | <0.001 |
| Oral corticosteroids (OCS) | 5.97 | 5.35-6.67 | <0.001 | - | - | - | - | - | - | 2.99 | 2.70-3.32 | <0.001 |
| Inhaled corticosteroids (ICS) | 6.32 | 5.69-7.02 | <0.001 | - | - | - | - | - | - | 1.85 | 1.62-2.11 | <0.001 |
| Long-acting beta-agonists (LABA) | 4.04 | 2.75-5.93 | <0.001 | - | - | - | - | - | - | 1.32 | 0.97-1.79 | 0.077 |
| Long-acting muscarinic antagonists (LAMA) ‡ | - | - | - | - | - | - | - | - | - | - | - | - |
| Leukotriene receptor antagonists (LKA) | 6.99 | 6.25-7.81 | <0.001 | - | - | - | - | - | - | 2.88 | 2.58-3.21 | <0.001 |
| Influenza vaccine | 1.46 | 1.33-1.60 | <0.001 | - | - | - | - | - | - | 0.95 | 0.86-1.04 | 0.240 |

*Adjusted for patient characteristics; ** Adjusted for patient characteristics and comorbidities; *** Adjusted for all covariates; † Medication use is recorded within 1 year of the index date;

‡ Number of cases too low to generate IRR

IRR, Incidence rate ratio

| **12-17 years**  ***Adjusted Incidence Rate Ratios (IRR) for hospital admissions among children aged 12-17 years***  **Supplementary table 2b** | | | | | | | | | | | | |
| --- | --- | --- | --- | --- | --- | --- | --- | --- | --- | --- | --- | --- |
|  | Univariable model | | | Multivariable models | | | | | | | | |
|  |  |  |  | Partially adjusted* | | | Partially adjusted** | | | Fully adjusted *** | | |
| Covariate | Unadjusted IRR | 95% CI | P-value | Adjusted IRR | 95% CI | P-value | Adjusted IRR | 95% CI | P-value | Adjusted IRR | 95% CI | P-value |
| Sex, n (%) |  |  |  |  |  |  |  |  |  |  |  |  |
| Male | 1 | - | - | 1 | - | - | 1 | - | - | 1 | - | - |
| Female | 1.97 | 1.71-2.27 | <0.001 | 1.98 | 1.71-2.28 | <0.001 | 1.99 | 1.72-2.29 | <0.001 | 1.83 | 1.61-2.08 | <0.001 |
| Ethnicity |  |  |  |  |  |  |  |  |  |  |  |  |
| White | 1 | - | - | 1 | - | - | 1 | - | - | 1 | - | - |
| Black | 1.87 | 1.32-2.65 | <0.001 | 2.08 | 1.48-2.93 | <0.001 | 1.76 | 1.25-2.47 | 0.001 | 1.74 | 1.31-2.32 | <0.001 |
| Mixed | 2.35 | 1.54-3.59 | <0.001 | 2.19 | 1.45-3.30 | <0.001 | 1.93 | 1.29-2.89 | 0.001 | 1.74 | 1.24-2.44 | 0.001 |
| Asian | 1.58 | 1.22-2.04 | <0.001 | 1.68 | 1.29-2.18 | <0.001 | 1.52 | 1.17-1.98 | 0.002 | 1.42 | 1.14-1.78 | 0.002 |
| Other | 1.22 | 0.62-2.37 | 0.565 | 1.52 | 0.80-2.90 | 0.206 | 1.29 | 0.68-2.45 | 0.439 | 2.01 | 1.21-3.34 | 0.007 |
| Missing | 0.74 | 0.63-0.86 | <0.001 | 0.89 | 0.74-1.06 | 0.180 | 0.88 | 0.74-1.05 | 0.163 | 0.96 | 0.82-1.12 | 0.595 |
| IMD score, n (%) |  |  |  |  |  |  |  |  |  |  |  |  |
| 1 – Least deprived | 1 | - | - | 1 | - | - | 1 | - | - | 1 | - | - |
| 2 | 1.22 | 0.97-1.55 | 0.095 | 1.15 | 0.90-1.49 | 0.267 | 1.16 | 0.91-1.49 | 0.237 | 1.15 | 0.92-1.44 | 0.226 |
| 3 | 1.47 | 1.16-1.86 | 0.001 | 1.27 | 0.98-1.63 | 0.068 | 1.29 | 1.00-1.65 | 0.049 | 1.19 | 0.94-1.49 | 0.145 |
| 4 | 1.60 | 1.27-2.01 | <0.001 | 1.46 | 1.14-1.87 | 0.003 | 1.52 | 1.19-1.94 | 0.001 | 1.28 | 1.03-1.60 | 0.029 |
| 5 – Most deprived | 1.74 | 1.39-2.16 | <0.001 | 1.50 | 1.17-1.92 | 0.001 | 1.54 | 1.21-1.96 | <0.001 | 1.52 | 1.22-1.89 | <0.001 |
| Missing ‡ | - | - | - | - | - | - | - | - | - | - | - | - |
| BMI, n (%) |  |  |  |  |  |  |  |  |  |  |  |  |
| Normal weight | 1 | - | - | 1 | - | - | 1 | - | - | 1 | - | - |
| Underweight | 0.53 | 0.42-0.66 | <0.001 | 0.55 | 0.43-0.69 | <0.001 | 0.57 | 0.45-0.72 | <0.001 | 0.76 | 0.62-0.95 | 0.014 |
| Overweight | 1.52 | 1.19-1.93 | 0.001 | 1.28 | 1.01-1.62 | 0.042 | 1.26 | 1.00-1.59 | 0.050 | 0.99 | 0.81-1.21 | 0.925 |
| Obese | 1.68 | 1.23-2.30 | 0.001 | 1.38 | 1.02-1.86 | 0.039 | 1.41 | 1.05-1.90 | 0.024 | 0.93 | 0.72-1.21 | 0.608 |
| Missing | 0.45 | 0.38-0.53 | <0.001 | 0.50 | 0.42-0.61 | <0.001 | 0.54 | 0.45-0.65 | <0.001 | 0.82 | 0.69-0.97 | 0.020 |
| Smoking status, n (%) |  |  |  |  |  |  |  |  |  |  |  |  |
| Never smoked | 1 | - | - | 1 | - | - | 1 | - | - | 1 | - | - |
| Current smoker | 1.04 | 0.83-1.32 | 0.714 | 1.20 | 0.95-1.52 | 0.125 | 1.23 | 0.97-1.55 | 0.082 | 1.12 | 0.91-1.39 | 0.280 |
| Former smoker | 1.88 | 1.51-2.35 | <0.001 | 1.82 | 1.46-2.26 | <0.001 | 1.68 | 1.35-2.09 | <0.001 | 1.26 | 1.05-1.53 | 0.015 |
| Missing | 0.39 | 0.32-0.46 | <0.001 | 0.57 | 0.47-0.69 | <0.001 | 0.61 | 0.50-0.74 | <0.001 | 1.05 | 0.88-1.27 | 0.581 |
| Comorbidities, n (%) |  |  |  |  |  |  |  |  |  |  |  |  |
| Allergies | 2.41 | 2.07-2.82 | <0.001 | - | - | - | 1.63 | 1.36-1.96 | <0.001 | 1.08 | 0.92-1.27 | 0.336 |
| Atopic eczema | 1.74 | 1.51-2.01 | <0.001 | - | - | - | 1.52 | 1.32-1.76 | <0.001 | 1.14 | 1.00-1.29 | 0.055 |
| Allergic rhinitis | 2.32 | 1.99-2.71 | <0.001 | - | - | - | 1.30 | 1.08-1.56 | 0.005 | 1.18 | 1.00-1.38 | 0.047 |
| Gastro-oesophageal reflux disease (GORD) | 2.29 | 1.56-3.38 | <0.001 | - | - | - | 1.59 | 1.09-2.31 | 0.015 | 1.14 | 0.81-1.61 | 0.438 |
| Chronic rhinosinusitis | 0.88 | 0.18-4.27 | 0.873 | - | - | - | 1.02 | 0.24-4.41 | 0.976 | 0.81 | 0.21-3.04 | 0.750 |
| Anxiety | 1.32 | 0.95-1.82 | 0.098 | - | - | - | 1.25 | 0.92-1.69 | 0.156 | 1.24 | 0.95-1.62 | 0.115 |
| Depression | 0.98 | 0.62-1.54 | 0.921 | - | - | - | 0.88 | 0.57-1.36 | 0.559 | 0.86 | 0.59-1.26 | 0.453 |
| Medication use, n (%)† |  |  |  |  |  |  |  |  |  |  |  |  |
| Number of SABA prescriptions within last year, n (%) |  |  |  |  |  |  |  |  |  |  |  |  |
| 0 | 1 | - | - | - | - | - | - | - | - | 1 | - | - |
| 1-3 | 3.18 | 2.72-3.70 | <0.001 | - | - | - | - | - | - | 1.36 | 1.10-1.67 | 0.004 |
| 4-6 | 10.56 | 8.64-12.89 | <0.001 | - | - | - | - | - | - | 2.38 | 1.84-3.07 | <0.001 |
| 7+ | 45.10 | 36.57-55.60 | <0.001 | - | - | - | - | - | - | 5.58 | 4.29-7.26 | <0.001 |
| Oral corticosteroids (OCS) | 19.78 | 16.19-24.17 | <0.001 | - | - | - | - | - | - | 4.09 | 3.44-4.85 | <0.001 |
| Inhaled corticosteroids (ICS) | 11.32 | 9.86-13.01 | <0.001 | - | - | - | - | - | - | 2.14 | 1.76-2.62 | <0.001 |
| Long-acting beta-agonists (LABA) | 12.47 | 6.95-22.40 | <0.001 | - | - | - | - | - | - | 2.06 | 1.45-2.93 | <0.001 |
| Long-acting muscarinic antagonists (LAMA) | 29.71 | 0.005-179775.3 | 0.445 | - | - | - | - | - | - | 5.00 | 0.14-179.64 | 0.378 |
| Leukotriene receptor antagonists (LKA) | 23.85 | 19.14-29.71 | <0.001 | - | - | - | - | - | - | 4.50 | 3.74-5.41 | <0.001 |
| Influenza vaccine | 4.03 | 3.43-4.73 | <0.001 | - | - | - | - | - | - | 1.07 | 0.92-1.23 | 0.385 |

*Adjusted for patient characteristics; ** Adjusted for patient characteristics and comorbidities; *** Adjusted for all covariates; † Medication use is recorded within 1 year of the index date;

‡ Number of cases too low to generate

IRR; IRR, Incidence rate ratio

| **18+ years**  ***Adjusted Incidence Rate Ratios (IRR) for hospital admissions among adults aged 18+ years***  **Supplementary table 2c** | | | | | | | | | | | | |
| --- | --- | --- | --- | --- | --- | --- | --- | --- | --- | --- | --- | --- |
|  | Univariable model | | | Multivariable models | | | | | | | | |
|  |  |  |  | Partially adjusted* | | | Partially adjusted** | | | Fully adjusted *** | | |
| Covariate | Unadjusted IRR | 95% CI | P-value | Adjusted IRR | 95% CI | P-value | Adjusted IRR | 95% CI | P-value | Adjusted IRR | 95% CI | P-value |
| Age (years) |  |  |  |  |  |  |  |  |  |  |  |  |
| 18-24 | 1 | - | - | 1 | - | - | 1 | - | - | 1 | - | - |
| 25-39 | 1.07 | 1.00-1.15 | 0.049 | 0.84 | 0.78-0.91 | <0.001 | 0.81 | 0.75-0.87 | <0.001 | 0.76 | 0.71-0.81 | <0.001 |
| 40-59 | 1.61 | 1.50-1.72 | <0.001 | 1.09 | 1.02-1.17 | 0.018 | 0.98 | 0.91-1.06 | 0.608 | 0.65 | 0.61-0.70 | <0.001 |
| 60-79 | 1.49 | 1.38-1.61 | <0.001 | 1.09 | 1.00-1.18 | 0.041 | 0.96 | 0.88-1.04 | 0.317 | 0.58 | 0.53-0.62 | <0.001 |
| 80+ | 2.16 | 1.92-2.43 | <0.001 | 1.88 | 1.68-2.11 | <0.001 | 1.70 | 1.52-1.91 | <0.001 | 1.06 | 0.96-1.18 | 0.257 |
| Sex, n (%) |  |  |  |  |  |  |  |  |  |  |  |  |
| Male | 1 | - | - | 1 | - | - | 1 | - | - | 1 | - | - |
| Female | 2.45 | 2.35-2.56 | <0.001 | 2.20 | 2.11-2.31 | <0.001 | 1.98 | 1.89-2.07 | <0.001 | 1.61 | 1.54-1.68 | <0.001 |
| Ethnicity |  |  |  |  |  |  |  |  |  |  |  |  |
| White | 1 | - | - | 1 | - | - | 1 | - | - | 1 | - | - |
| Black | 1.35 | 1.19-1.54 | <0.001 | 1.19 | 1.04-1.36 | 0.010 | 1.24 | 1.09-1.42 | 0.001 | 1.46 | 1.30-1.64 | <0.001 |
| Mixed | 1.05 | 0.89-1.25 | 0.547 | 1.07 | 0.91-1.27 | 0.407 | 1.09 | 0.92-1.29 | 0.315 | 1.25 | 1.07-1.45 | 0.004 |
| Asian | 1.63 | 1.50-1.77 | <0.001 | 1.69 | 1.54-1.85 | <0.001 | 1.65 | 1.51-1.81 | <0.001 | 1.60 | 1.48-1.74 | <0.001 |
| Other | 1.46 | 1.19-1.78 | <0.001 | 1.36 | 1.12-1.66 | 0.002 | 1.40 | 1.15-1.70 | 0.001 | 1.17 | 0.98-1.41 | 0.085 |
| Missing | 0.68 | 0.65-0.72 | <0.001 | 0.80 | 0.75-0.85 | <0.001 | 0.80 | 0.76-0.85 | <0.001 | 0.90 | 0.85-0.95 | <0.001 |
| IMD score, n (%) |  |  |  |  |  |  |  |  |  |  |  |  |
| 1 – Least deprived | 1 | - | - | 1 | - | - | 1 | - | - | 1 | - | - |
| 2 | 1.10 | 1.03-1.18 | 0.005 | 1.10 | 1.02-1.18 | 0.014 | 1.11 | 1.03-1.19 | 0.007 | 1.08 | 1.00-1.15 | 0.036 |
| 3 | 1.18 | 1.10-1.26 | <0.001 | 1.21 | 1.12-1.31 | <0.001 | 1.21 | 1.13-1.31 | <0.001 | 1.17 | 1.09-1.25 | <0.001 |
| 4 | 1.56 | 1,46-1.66 | <0.001 | 1.53 | 1.42-1.65 | <0.001 | 1.49 | 1.38-1.60 | <0.001 | 1.38 | 1.28-1.48 | <0.001 |
| 5 – Most deprived | 1.88 | 1.76-2.01 | <0.001 | 1.69 | 1.56-1.83 | <0.001 | 1.64 | 1.52-1.78 | <0.001 | 1.43 | 1.33-1.54 | <0.001 |
| Missing | 0.49 | 0.21-1.16 | 0.106 | 0.59 | 0.24-1.44 | 0.245 | 0.67 | 0.28-1.61 | 0.370 | 0.75 | 0.33-1.71 | 0.499 |
| BMI, n (%) |  |  |  |  |  |  |  |  |  |  |  |  |
| Normal weight | 1 | - | - | 1 | - | - | 1 | - | - | 1 | - | - |
| Underweight | 0.98 | 0.85-1.12 | 0.743 | 1.03 | 0.90-1.19 | 0.636 | 1.08 | 0.94-1.24 | 0.305 | 1.21 | 1.06-1.38 | 0.005 |
| Overweight | 1.34 | 1.27-1.42 | <0.001 | 1.31 | 1.23-1.38 | <0.001 | 1.27 | 1.20-1.34 | <0.001 | 1.12 | 1.06-1.18 | <0.001 |
| Obese | 2.53 | 2.40-2.67 | <0.001 | 2.21 | 2.09-2.33 | <0.001 | 2.08 | 1.96-2.19 | <0.001 | 1.51 | 1.43-1.59 | <0.001 |
| Missing | 0.62 | 0.57-0.68 | <0.001 | 0.88 | 0.81-0.97 | 0.009 | 0.93 | 0.85-1.02 | 0.148 | 1.17 | 1.07-1.28 | 0.001 |
| Smoking status, n (%) |  |  |  |  |  |  |  |  |  |  |  |  |
| Never smoked | 1 | - | - | 1 | - | - | 1 | - | - | 1 | - | - |
| Current smoker | 1.18 | 1.12-1.25 | <0.001 | 1.28 | 1.21-1.36 | <0.001 | 1.22 | 1.15-1.29 | <0.001 | 1.19 | 1.12-1.25 | <0.001 |
| Former smoker | 1.39 | 1.32-1.46 | <0.001 | 1.26 | 1.19-1.33 | <0.001 | 1.17 | 1.11-1.24 | <0.001 | 1.03 | 0.98-1.08 | 0.219 |
| Missing | 0.21 | 0.16-0.28 | <0.001 | 0.46 | 0.34-0.62 | <0.001 | 0.51 | 0.38-0.68 | <0.001 | 0.82 | 0.62-1.09 | 0.174 |
| Comorbidities, n (%) |  |  |  |  |  |  |  |  |  |  |  |  |
| Allergies | 1.72 | 1.64-1.80 | <0.001 | - | - | - | 1.46 | 1.38-1.55 | <0.001 | 1.20 | 1.14-1.27 | <0.001 |
| Atopic eczema | 1.28 | 1.22-1.34 | <0.001 | - | - | - | 1.21 | 1.15-1.26 | <0.001 | 1.02 | 0.98-1.07 | 0.280 |
| Allergic rhinitis | 1.40 | 1.34-1.47 | <0.001 | - | - | - | 1.02 | 0.96-1.08 | 0.568 | 0.90 | 0.85-0.95 | <0.001 |
| Gastro-oesophageal reflux disease (GORD) | 2.01 | 1.88-2.14 | <0.001 | - | - | - | 1.49 | 1.40-1.58 | <0.001 | 1.14 | 1.07-1.20 | <0.001 |
| Chronic rhinosinusitis | 2.06 | 1.81-2.33 | <0.001 | - | - | - | 1.69 | 1.50-1.91 | <0.001 | 1.21 | 1.08-1.35 | 0.001 |
| Anxiety | 1.68 | 1.60-1.76 | <0.001 | - | - | - | 1.16 | 1.10-1.22 | <0.001 | 1.07 | 1.02-1.12 | 0.010 |
| Depression | 2.12 | 2.03-2.22 | <0.001 | - | - | - | 1.51 | 1.43-1.58 | <0.001 | 1.28 | 1.22-1.34 | <0.001 |
| Medication use, n (%)† |  |  |  |  |  |  |  |  |  |  |  |  |
| Number of SABA prescriptions within last year, n (%) |  |  |  |  |  |  |  |  |  |  |  |  |
| 0 | 1 | - | - | - | - | - | - | - | - | 1 | - | - |
| 1-3 | 3.73 | 3.55-3.91 | <0.001 | - | - | - | - | - | - | 1.47 | 1.39-1.56 | <0.001 |
| 4-6 | 7.69 | 7.22-8.20 | <0.001 | - | - | - | - | - | - | 1.92 | 1.79-2.06 | <0.001 |
| 7+ | 14.02 | 13.19-14.91 | <0.001 | - | - | - | - | - | - | 2.52 | 2.35-2.70 | <0.001 |
| Oral corticosteroids (OCS) | 11.01 | 10.51-11.53 | <0.001 | - | - | - | - | - | - | 3.48 | 3.32-3.64 | <0.001 |
| Inhaled corticosteroids (ICS) | 8.01 | 7.67-8.37 | <0.001 | - | - | - | - | - | - | 2.21 | 2.09-2.34 | <0.001 |
| Long-acting beta-agonists (LABA) | 4.71 | 4.36-5.09 | <0.001 | - | - | - | - | - | - | 1.37 | 1.29-1.46 | <0.001 |
| Long-acting muscarinic antagonists (LAMA) | 32.77 | 27.44-39.13 | <0.001 | - | - | - | - | - | - | 5.07 | 4.50-5.71 | <0.001 |
| Leukotriene receptor antagonists (LKA) | 15.72 | 14.49-17.06 | <0.001 | - | - | - | - | - | - | 3.19 | 2.98-3.41 | <0.001 |
| Influenza vaccine | 2.91 | 2.79-3.03 | <0.001 | - | - | - | - | - | - | 1.09 | 1.04-1.14 | <0.001 |

*Adjusted for patient characteristics; ** Adjusted for patient characteristics and comorbidities; *** Adjusted for all covariates; † Medication use is recorded within 1 year of the index date;

IRR, Incidence rate ratio

| **Risk factor** | **Age group** | | |
| --- | --- | --- | --- |
|  | **5-11 years** | **12-17 years** | **18+ years** |
|  | *PAF (%) (95% CI)** | *PAF (%) (95% CI)** | *PAF (%) (95% CI)** |
| Obesity | - | - | 23.3 (20.5-26.1) |
| Current smoker | - | - | 4.3 (3.0-5.7) |
| Current/ex-smoker | - | 6.8 (0.9-12.3) | - |
| Allergies | 6.8 (3.6-9.9) | - | 6.2 (4.4-8.0) |
| Atopic eczema | 11.4 (6.8-15.8) | - | - |
| Allergic rhinitis | - | 6.3 (-0.03-12.3) | - |
| GORD | - | - | 2.3 (1.2-3.4) |
| Chronic rhinosinusitis | - | - | 0.8 (0.3-1.3) |
| Anxiety | - | - | 2.0 (0.5-3.6) |
| Depression | - | - | 11.1 (9.1-13.1) |

*Adjusted for all covariates

GORD=gastroesophageal reflux disease, PAF=population attributable risk fraction

***Population attributable risk fraction of selected risk factors for asthma-related hospital admissions***

**Supplementary table 3**

| **5-11 years**  ***Adjusted Incidence Rate Ratios (IRR) for ICU admissions among children aged 5-11 years***  **Supplementary table 4a** | | | | | | | | | | | | |
| --- | --- | --- | --- | --- | --- | --- | --- | --- | --- | --- | --- | --- |
|  | Univariable model | | | Multivariable models | | | | | | | | |
|  |  |  |  | Partially adjusted* | | | Partially adjusted** | | | Fully adjusted *** | | |
| Covariate | Unadjusted IRR | 95% CI | P-value | Adjusted IRR | 95% CI | P-value | Adjusted IRR | 95% CI | P-value | Adjusted IRR | 95% CI | P-value |
| Sex, n (%) |  |  |  |  |  |  |  |  |  |  |  |  |
| Male | 1 | - | - | 1 | - | - | 1 | - | - | 1 | - | - |
| Female | 1.17 | 0.82-1.65 | 0.384 | 1.15 | 0.81-1.63 | 0.421 | 1.15 | 0.81-1.64 | 0.421 | 1.20 | 0.85-1.70 | 0.306 |
| Ethnicity |  |  |  |  |  |  |  |  |  |  |  |  |
| White | 1 | - | - | 1 | - | - | 1 | - | - | 1 | - | - |
| Black | 5.10 | 3.07-8.49 | <0.001 | 3.94 | 2.28-6.78 | <0.001 | 3.78 | 2.18-6.55 | <0.001 | 4.07 | 2.35-7.05 | <0.001 |
| Mixed | 2.79 | 1.39-5.58 | 0.004 | 2.49 | 1.23-5.04 | 0.011 | 2.42 | 1.20-4.91 | 0.014 | 2.49 | 1.23-5.05 | 0.011 |
| Asian | 1.53 | 0.87-2.70 | 0.137 | 1.35 | 0.75-2.42 | 0.316 | 1.28 | 0.71-2.32 | 0.406 | 1.17 | 0.65-2.12 | 0.598 |
| Other | 1.37 | 0.33-5.78 | 0.666 | 1.22 | 0.29-5.19 | 0.787 | 1.20 | 0.28-5.09 | 0.808 | 1.26 | 0.30-5.35 | 0.750 |
| Missing | 1.55 | 0.97-2.46 | 0.066 | 1.56 | 0.97-2.51 | 0.065 | 1.56 | 0.97-2.51 | 0.067 | 1.56 | 0.97-2.52 | 0.068 |
| IMD score, n (%) |  |  |  |  |  |  |  |  |  |  |  |  |
| 1 – Least deprived | 1 | - | - | 1 | - | - | 1 | - | - | 1 | - | - |
| 2 | 1.52 | 0.74-3.13 | 0.259 | 1.47 | 0.71-3.05 | 0.301 | 1.48 | 0.71-3.07 | 0.295 | 1.41 | 0.68-2.93 | 0.358 |
| 3 | 1.87 | 0.93-3.75 | 0.079 | 1.71 | 0.84-3.47 | 0.137 | 1.72 | 0.85-3.49 | 0.134 | 1.59 | 0.78-3.22 | 0.202 |
| 4 | 2.28 | 1.18-4.40 | 0.014 | 1.78 | 0.90-3.51 | 0.096 | 1.80 | 0.91-3.55 | 0.090 | 1.63 | 0.82-3.22 | 0.160 |
| 5 – Most deprived | 2.95 | 1.59-5.48 | 0.001 | 2.31 | 1.21-4.42 | 0.012 | 2.33 | 1.22-4.46 | 0.011 | 1.98 | 1.03-3.79 | 0.041 |
| Missing ‡ | - | - | - | - | - | - | - | - | - | - | - | - |
| BMI, n (%) |  |  |  |  |  |  |  |  |  |  |  |  |
| Normal weight | 1 | - | - | 1 | - | - | 1 | - | - | 1 | - | - |
| Underweight | 0.96 | 0.42-2.18 | 0.927 | 0.97 | 0.43-2.21 | 0.942 | 0.97 | 0.43-2.21 | 0.940 | 1.08 | 0.47-2.45 | 0.858 |
| Overweight | 1.78 | 0.99-3.20 | 0.053 | 1.58 | 0.88-2.85 | 0.126 | 1.58 | 0.88-2.84 | 0.129 | 1.53 | 0.85-2.74 | 0.158 |
| Obese | 0.93 | 0.33-2.66 | 0.897 | 0.79 | 0.27-2.25 | 0.653 | 0.79 | 0.29-2.28 | 0.667 | 0.67 | 0.23-1.90 | 0.448 |
| Missing | 1.15 | 0.77-1.71 | 0.488 | 1.12 | 0.75-1.68 | 0.579 | 1.14 | 0.76-1.71 | 0.531 | 1.47 | 0.97-2.21 | 0.067 |
| Comorbidities, n (%) |  |  |  |  |  |  |  |  |  |  |  |  |
| Allergies | 1.35 | 0.91-2.01 | 0.140 | - | - | - | 1.37 | 0.86-2.18 | 0.182 | 1.16 | 0.74-1.83 | 0.520 |
| Atopic eczema | 1.32 | 0.94-1.87 | 0.112 | - | - | - | 1.24 | 0.87-1.77 | 0.224 | 1.02 | 0.72-1.46 | 0.894 |
| Allergic rhinitis | 1.03 | 0.65-1.63 | 0.916 | - | - | - | 0.75 | 0.44-1.28 | 0.294 | 0.67 | 0.40-1.14 | 0.141 |
| Gastro-oesophageal reflux disease (GORD) | 0.62 | 0.25-1.54 | 0.304 | - | - | - | 0.69 | 0.28-1.72 | 0.425 | 0.57 | 0.23-1.43 | 0.233 |
| Chronic rhinosinusitis ‡ | - | - | - | - | - | - | - | - | - | - | - | - |
| Anxiety | 0.70 | 0.09-5.21 | 0.728 | - | - | - | 0.80 | 0.11-5.95 | 0.824 | 0.76 | 0.10-5.75 | 0.791 |
| Depression | 0.72 | 0.10-5.38 | 0.751 | - | - | - | 0.93 | 0.12-7.14 | 0.946 | 0.79 | 0.10-6.11 | 0.825 |
| Medication use, n (%) † |  |  |  |  |  |  |  |  |  |  |  |  |
| Number of SABA prescriptions within last year, n (%) |  |  |  |  |  |  |  |  |  |  |  |  |
| 0 | 1 | - | - | - | - | - | - | - | - | 1 | - | - |
| 1-3 | 3.19 | 1.65-6.17 | 0.001 | - | - | - | - | - | - | 2.13 | 0.98-4.61 | 0.056 |
| 4-6 | 11.12 | 5.71-21.67 | <0.001 | - | - | - | - | - | - | 5.21 | 2.26-12.01 | <0.001 |
| 7+ | 18.27 | 9.18-36.36 | <0.001 | - | - | - | - | - | - | 6.97 | 2.91-16.68 | <0.001 |
| Oral corticosteroids (OCS) | 4.28 | 2.97-6.16 | <0.001 | - | - | - | - | - | - | 1.98 | 1.34-2.92 | 0.001 |
| Inhaled corticosteroids (ICS) | 4.27 | 2.71-6.72 | <0.001 | - | - | - | - | - | - | 1.31 | 0.73-2.34 | 0.371 |
| Long-acting beta-agonists (LABA) | 2.89 | 0.99-8.42 | 0.052 | - | - | - | - | - | - | 1.23 | 0.43-3.53 | 0.699 |
| Long-acting muscarinic antagonists (LAMA) ‡ | - | - | - | - | - | - | - | - | - | - | - | - |
| Leukotriene receptor antagonists (LKA) | 6.00 | 4.21-8.54 | <0.001 | - | - | - | - | - | - | 2.86 | 1.93-4.24 | <0.001 |
| Influenza vaccine | 1.42 | 1.00-2.01 | 0.047 | - | - | - | - | - | - | 1.00 | 0.70-1.44 | 0.988 |

*Adjusted for patient characteristics; ** Adjusted for patient characteristics and comorbidities; *** Adjusted for all covariates; † Medication use is recorded within 1 year of the index date;

‡ Number of cases too low to generate IRR

IRR, Incidence rate ratio

| **12-17 years**  ***Adjusted Incidence Rate Ratios (IRR) for ICU admissions among children aged 12-17 years***  **Supplementary table 4b** | | | | | | | | | | | | |
| --- | --- | --- | --- | --- | --- | --- | --- | --- | --- | --- | --- | --- |
|  | Univariable model | | | Multivariable models | | | | | | | | |
|  |  |  |  | Partially adjusted* | | | Partially adjusted** | | | Fully adjusted *** | | |
| Covariate | Unadjusted IRR | 95% CI | P-value | Adjusted IRR | 95% CI | P-value | Adjusted IRR | 95% CI | P-value | Adjusted IRR | 95% CI | P-value |
| Sex, n (%) |  |  |  |  |  |  |  |  |  |  |  |  |
| Male | 1 | - | - | 1 | - | - | 1 | - | - | 1 | - | - |
| Female | 1.57 | 1.02-2.41 | 0.038 | 1.55 | 1.01-2.38 | 0.046 | 1.59 | 1.03-2.46 | 0.035 | 1.54 | 1.00-2.36 | 0.048 |
| Ethnicity |  |  |  |  |  |  |  |  |  |  |  |  |
| White | 1 | - | - | 1 | - | - | 1 | - | - | 1 | - | - |
| Black | 3.66 | 1.70-7.85 | 0.001 | 3.18 | 1.46-6.94 | 0.004 | 2.72 | 1.24-5.98 | 0.012 | 3.51 | 1.62-7.59 | 0.001 |
| Mixed | 2.85 | 1.04-7.79 | 0.041 | 2.65 | 0.96-7.33 | 0.060 | 2.33 | 0.85-6.44 | 0.102 | 2.43 | 0.90-6.54 | 0.080 |
| Asian | 2.11 | 1.06-4.22 | 0.034 | 1.82 | 0.90-3.67 | 0.097 | 1.59 | 0.78-3.22 | 0.199 | 1.46 | 0.73-2.93 | 0.287 |
| Other ‡ | - | - | - | - | - | - | - | - | - | - | - | - |
| Missing | 1.26 | 0.77-2.09 | 0.359 | 1.62 | 0.97-2.71 | 0.066 | 1.60 | 0.95-2.68 | 0.074 | 1.69 | 1.01-2.82 | 0.046 |
| IMD score, n (%) |  |  |  |  |  |  |  |  |  |  |  |  |
| 1 – Least deprived | 1 | - | - | 1 | - | - | 1 | - | - | 1 | - | - |
| 2 | 2.35 | 0.94-5.88 | 0.068 | 2.33 | 0.93-5.84 | 0.072 | 2.34 | 0.93-5.86 | 0.070 | 2.08 | 0.83-5.19 | 0.117 |
| 3 | 2.89 | 1.18-7.07 | 0.020 | 2.68 | 1.09-6.60 | 0.032 | 2.70 | 1.10-6.64 | 0.030 | 2.32 | 0.95-5.66 | 0.066 |
| 4 | 3.39 | 1.43-8.03 | 0.005 | 3.01 | 1.25-7.22 | 0.014 | 3.06 | 1.28-7.34 | 0.012 | 2.42 | 1.01-5.77 | 0.046 |
| 5 – Most deprived | 3.35 | 1.44-7.81 | 0.005 | 2.73 | 1.15-6.51 | 0.023 | 2.75 | 1.16-6.56 | 0.022 | 2.09 | 0.88-4.98 | 0.096 |
| Missing ‡ | - | - | - | - | - | - | - | - | - | - | - | - |
| BMI, n (%) |  |  |  |  |  |  |  |  |  |  |  |  |
| Normal weight | 1 | - | - | 1 | - | - | 1 | - | - | 1 | - | - |
| Underweight | 0.75 | 0.38-1.48 | 0.409 | 0.77 | 0.39-1.53 | 0.458 | 0.79 | 0.40-1.56 | 0.493 | 1.04 | 0.53-2.06 | 0.900 |
| Overweight | 1.28 | 0.67-2.44 | 0.458 | 1.18 | 0.61-2.26 | 0.623 | 1.12 | 0.58-2.15 | 0.732 | 0.91 | 0.48-1.73 | 0.781 |
| Obese | 1.42 | 0.63-3.20 | 0.398 | 1.20 | 0.53-2.74 | 0.658 | 1.18 | 0.52-2.68 | 0.694 | 0.85 | 0.38-1.91 | 0.701 |
| Missing | 0.44 | 0.25-0.78 | 0.004 | 0.54 | 0.30-0.97 | 0.038 | 0.58 | 0.32-1.05 | 0.073 | 0.86 | 0.48-1.54 | 0.618 |
| Smoking status, n (%) |  |  |  |  |  |  |  |  |  |  |  |  |
| Never smoked | 1 | - | - | 1 | - | - | 1 | - | - | 1 | - | - |
| Current smoker | 0.88 | 0.42-1.83 | 0.731 | 0.83 | 0.40-1.74 | 0.620 | 0.83 | 0.40-1.73 | 0.617 | 0.83 | 0.40-1.73 | 0.616 |
| Former smoker | 1.73 | 0.98-3.05 | 0.060 | 1.57 | 0.88-2.80 | 0.130 | 1.47 | 0.82-2.63 | 0.194 | 1.14 | 0.65-2.01 | 0.655 |
| Missing | 0.45 | 0.24-0.86 | 0.016 | 0.55 | 0.28-1.06 | 0.074 | 0.60 | 0.31-1.17 | 0.133 | 1.09 | 0.55-2.13 | 0.807 |
| Comorbidities, n (%) |  |  |  |  |  |  |  |  |  |  |  |  |
| Allergies | 2.64 | 1.72-4.04 | <0.001 | - | - | - | 1.91 | 1.12-3.25 | 0.017 | 1.51 | 0.89-2.54 | 0.124 |
| Atopic eczema | 1.63 | 1.06-2.50 | 0.027 | - | - | - | 1.29 | 0.83-2.00 | 0.261 | 0.86 | 0.56-1.35 | 0.521 |
| Allergic rhinitis | 2.13 | 1.38-3.29 | 0.001 | - | - | - | 1.25 | 0.73-2.13 | 0.421 | 1.05 | 0.62-1.77 | 0.854 |
| Gastro-oesophageal reflux disease (GORD) | 1.45 | 0.50-4.19 | 0.497 | - | - | - | 1.44 | 0.50-4.19 | 0.502 | 1.14 | 0.40-3.27 | 0.806 |
| Chronic rhinosinusitis ‡ | - | - | - | - | - | - | - | - | - | - | - | - |
| Anxiety | 1.13 | 0.44-2.92 | 0.797 | - | - | - | 0.87 | 0.33-2.33 | 0.789 | 0.94 | 0.36-2.46 | 0.906 |
| Depression | 2.65 | 1.07-6.59 | 0.036 | - | - | - | 2.08 | 0.82-5.32 | 0.125 | 1.90 | 0.77-4.70 | 0.166 |
| Medication use, n (%) † |  |  |  |  |  |  |  |  |  |  |  |  |
| Number of SABA prescriptions within last year, n (%) |  |  |  |  |  |  |  |  |  |  |  |  |
| 0 | 1 | - | - | - | - | - | - | - | - | 1 | - | - |
| 1-3 | 6.18 | 2.50-15.26 | <0.001 | - | - | - | - | - | - | 1.71 | 0.53-5.50 | 0.369 |
| 4-6 | 20.14 | 7.90-51.37 | <0.001 | - | - | - | - | - | - | 3.03 | 0.86-10.64 | 0.083 |
| 7+ | 79.83 | 33.85-188.26 | <0.001 | - | - | - | - | - | - | 8.44 | 2.49-28.56 | 0.001 |
| Oral corticosteroids (OCS) | 17.15 | 11.19-26.29 | <0.001 | - | - | - | - | - | - | 4.06 | 2.56-6.45 | <0.001 |
| Inhaled corticosteroids (ICS) | 20.01 | 9.64-41.52 | <0.001 | - | - | - | - | - | - | 3.95 | 1.40-11.17 | 0.009 |
| Long-acting beta-agonists (LABA) | 8.70 | 3.60-20.99 | <0.001 | - | - | - | - | - | - | 1.72 | 0.78-3.79 | 0.180 |
| Long-acting muscarinic antagonists (LAMA) ‡ | - | - | - | - | - | - | - | - | - | - | - | - |
| Leukotriene receptor antagonists (LKA) | 16.45 | 10.61-25.50 | <0.001 | - | - | - | - | - | - | 2.77 | 1.71-4.49 | <0.001 |
| Influenza vaccine | 3.06 | 1.98-4.74 | <0.001 | - | - | - | - | - | - | 0.89 | 0.57-1.39 | 0.616 |

*Adjusted for patient characteristics; ** Adjusted for patient characteristics and comorbidities; *** Adjusted for all covariates; † Medication use is recorded within 1 year of the index date;

‡ Number of cases too low to generate IRR

IRR, Incidence rate ratio

| **18+ years** | | | | | | | | | | | | |
| --- | --- | --- | --- | --- | --- | --- | --- | --- | --- | --- | --- | --- |
|  | Univariable model | | | Multivariable models | | | | | | | | |
|  |  |  |  | Partially adjusted* | | | Partially adjusted** | | | Fully adjusted *** | | |
| Covariate  ***Adjusted Incidence Rate Ratios (IRR) for ICU admissions among adults aged 18+ years***  **Supplementary table 4c** | Unadjusted IRR | 95% CI | P-value | Adjusted IRR | 95% CI | P-value | Adjusted IRR | 95% CI | P-value | Adjusted IRR | 95% CI | P-value |
| Age (years) |  |  |  |  |  |  |  |  |  |  |  |  |
| 18-24 | 1 | - | - | 1 | - | - | 1 | - | - | 1 | - | - |
| 25-39 | 0.92 | 0.68-1.24 | 0.578 | 0.77 | 0.56-1.04 | 0.090 | 0.70 | 0.52-0.96 | 0.026 | 0.66 | 0.48-0.90 | 0.009 |
| 40-59 | 1.10 | 0.82-1.47 | 0.516 | 0.83 | 0.61-1.14 | 0.251 | 0.70 | 0.51-0.96 | 0.028 | 0.44 | 0.32-0.60 | <0.001 |
| 60-79 | 0.93 | 0.66-1.31 | 0.668 | 0.71 | 0.49-1.02 | 0.061 | 0.56 | 0.39-0.82 | 0.003 | 0.29 | 0.20-0.43 | <0.001 |
| 80+ | 0.62 | 0.32-1.21 | 0.162 | 0.51 | 0.26-0.99 | 0.047 | 0.38 | 0.19-0.75 | 0.005 | 0.19 | 0.10-0.38 | <0.001 |
| Sex, n (%) |  |  |  |  |  |  |  |  |  |  |  |  |
| Male | 1 | - | - | 1 | - | - | 1 | - | - | 1 | - | - |
| Female | 2.22 | 1.80-2.73 | <0.001 | 2.06 | 1.66-2.55 | <0.001 | 1.87 | 1.51-2.32 | <0.001 | 1.46 | 1.17-1.81 | 0.001 |
| Ethnicity |  |  |  |  |  |  |  |  |  |  |  |  |
| White | 1 | - | - | 1 | - | - | 1 | - | - | 1 | - | - |
| Black | 2.34 | 1.52-3.60 | <0.001 | 1.97 | 1.27-3.05 | 0.002 | 2.15 | 1.38-3.33 | 0.001 | 2.69 | 1.75-4.13 | <0.001 |
| Mixed | 1.66 | 0.89-3.11 | 0.114 | 1.51 | 0.81-2.83 | 0.198 | 1.55 | 0.83-2.91 | 0.169 | 2.03 | 1.10-3.74 | 0.023 |
| Asian | 1.44 | 1.01-2.05 | 0.044 | 1.51 | 1.05-2.16 | 0.025 | 1.53 | 1.07-2.20 | 0.020 | 1.46 | 1.02-2.07 | 0.038 |
| Other | 0.87 | 0.31-2.45 | 0.793 | 0.83 | 0.30-2.34 | 0.725 | 0.87 | 0.31-2.45 | 0.793 | 0.94 | 0.34-2.65 | 0.912 |
| Missing | 0.66 | 0.51-0.86 | 0.002 | 0.74 | 0.56-0.97 | 0.029 | 0.76 | 0.58-1.00 | 0.048 | 0.87 | 0.66-1.14 | 0.301 |
| IMD score, n (%) |  |  |  |  |  |  |  |  |  |  |  |  |
| 1 – Least deprived | 1 | - | - | 1 | - | - | 1 | - | - | 1 | - | - |
| 2 | 1.51 | 1.06-2.14 | 0.022 | 1.42 | 1.00-2.02 | 0.050 | 1.41 | 0.99-2.00 | 0.058 | 1.34 | 0.94-1.90 | 0.107 |
| 3 | 1.73 | 1.23-2.45 | 0.002 | 1.55 | 1.10-2.20 | 0.013 | 1.52 | 1.07-2.15 | 0.019 | 1.37 | 0.97-1.93 | 0.079 |
| 4 | 2.19 | 1.58-3.06 | <0.001 | 1.79 | 1.28-2.50 | 0.001 | 1.70 | 1.21-2.37 | 0.002 | 1.41 | 1.00-1.98 | 0.048 |
| 5 – Most deprived | 2.60 | 1.88-3.60 | <0.001 | 1.94 | 1.39-2.70 | <0.001 | 1.79 | 1.28-2.50 | 0.001 | 1.35 | 0.96-1.89 | 0.082 |
| Missing | 3.81 | 0.42-34.28 | 0.233 | 4.31 | 0.48-38.99 | 0.194 | 4.04 | 0.45-36.04 | 0.211 | 3.56 | 0.44-28.98 | 0.235 |
| BMI, n (%) |  |  |  |  |  |  |  |  |  |  |  |  |
| Normal weight | 1 | - | - | 1 | - | - | 1 | - | - | 1 | - | - |
| Underweight | 0.85 | 0.42-1.71 | 0.642 | 0.84 | 0.41-1.71 | 0.629 | 0.85 | 0.42-1.72 | 0.643 | 0.94 | 0.46-1.90 | 0.860 |
| Overweight | 0.95 | 0.72-1.26 | 0.718 | 1.00 | 0.75-1.32 | 0.975 | 0.96 | 0.73-1.28 | 0.798 | 0.83 | 0.62-1.00 | 0.194 |
| Obese | 2.27 | 1.78-2.88 | <0.001 | 2.06 | 1.61-2.64 | <0.001 | 1.88 | 1.46-2.41 | <0.001 | 1.25 | 0.97-1.61 | 0.080 |
| Missing | 1.01 | 0.70-1.46 | 0.944 | 1.36 | 0.93-2.00 | 0.117 | 1.41 | 0.96-2.08 | 0.078 | 1.76 | 1.20-2.60 | 0.004 |
| Smoking status, n (%) |  |  |  |  |  |  |  |  |  |  |  |  |
| Never smoked | 1 | - | - | 1 | - | - | 1 | - | - | 1 | - | - |
| Current smoker | 1.69 | 1.31-2.17 | <0.001 | 1.60 | 1.24-2.07 | <0.001 | 1.45 | 1.12-1.88 | 0.004 | 1.28 | 0.99-1.66 | 0.063 |
| Former smoker | 1.46 | 1.14-1.85 | 0.002 | 1.36 | 1.06-1.74 | 0.015 | 1.27 | 0.99-1.62 | 0.063 | 1.10 | 0.85-1.41 | 0.470 |
| Missing | 0.55 | 0.17-1.78 | 0.320 | 0.72 | 0.22-2.38 | 0.590 | 0.74 | 0.22-2.45 | 0.621 | 1.22 | 0.37-3.99 | 0.747 |
| Comorbidities, n (%) |  |  |  |  |  |  |  |  |  |  |  |  |
| Allergies | 1.33 | 1.09-1.64 | 0.006 | - | - | - | 1.32 | 1.02-1.70 | 0.038 | 1.13 | 0.88-1.47 | 0.334 |
| Atopic eczema | 1.07 | 0.86-1.32 | 0.541 | - | - | - | 0.99 | 0.80-1.23 | 0.929 | 0.88 | 0.71-1.10 | 0.261 |
| Allergic rhinitis | 1.11 | 0.90-1.37 | 0.314 | - | - | - | 0.89 | 0.69-1.16 | 0.405 | 0.78 | 0.60-1.01 | 0.063 |
| Gastro-oesophageal reflux disease (GORD) | 1.65 | 1.26-2.15 | <0.001 | - | - | - | 1.39 | 1.05-1.83 | 0.020 | 1.11 | 0.84-1.46 | 0.458 |
| Chronic rhinosinusitis | 0.64 | 0.29-1.39 | 0.257 | - | - | - | 0.56 | 0.25-1.21 | 0.140 | 0.43 | 0.20-0.94 | 0.035 |
| Anxiety | 1.65 | 1.34-2.03 | <0.001 | - | - | - | 1.03 | 0.82-1.30 | 0.785 | 0.94 | 0.75-1.19 | 0.627 |
| Depression | 2.38 | 1.97-2.88 | <0.001 | - | - | - | 1.94 | 1.56-2.42 | <0.001 | 1.60 | 1.28-2.00 | <0.001 |
| Medication use, n (%)† |  |  |  |  |  |  |  |  |  |  |  |  |
| Number of SABA prescriptions within last year, n (%) |  |  |  |  |  |  |  |  |  |  |  |  |
| 0 | 1 | - | - | - | - | - | - | - | - | 1 | - | - |
| 1-3 | 3.96 | 2.99-5.24 | <0.001 | - | - | - | - | - | - | 1.89 | 1.34-2.67 | <0.001 |
| 4-6 | 7.85 | 5.72-10.76 | <0.001 | - | - | - | - | - | - | 2.58 | 1.73-3.85 | <0.001 |
| 7+ | 18.33 | 13.96-24.06 | <0.001 | - | - | - | - | - | - | 4.39 | 3.02-6.37 | <0.001 |
| Oral corticosteroids (OCS) | 10.17 | 8.40-12.30 | <0.001 | - | - | - | - | - | - | 3.33 | 2.68-4.14 | <0.001 |
| Inhaled corticosteroids (ICS) | 7.13 | 5.63-9.02 | <0.001 | - | - | - | - | - | - | 1.90 | 1.36-2.67 | <0.001 |
| Long-acting beta-agonists (LABA) | 4.77 | 3.70-6.15 | <0.001 | - | - | - | - | - | - | 1.31 | 1.01-1.70 | 0.042 |
| Long-acting muscarinic antagonists (LAMA) | 27.51 | 19.74-38.36 | <0.001 | - | - | - | - | - | - | 4.28 | 3.05-6.01 | <0.001 |
| Leukotriene receptor antagonists (LKA) | 14.89 | 11.93-18.60 | <0.001 | - | - | - | - | - | - | 3.17 | 2.46-4.07 | <0.001 |
| Influenza vaccine | 2.55 | 2.11-3.09 | <0.001 | - | - | - | - | - | - | 1.10 | 0.88-1.36 | 0.405 |

*Adjusted for patient characteristics; ** Adjusted for patient characteristics and comorbidities; *** Adjusted for all covariates; † Medication use is recorded within 1 year of the index date;

IRR, Incidence rate ratio

**Supplementary figures**

***Supplementary figure 1***

**Bar chart showing the distribution of hospital admission frequency among all hospitalised patients**

Percentage of patients

| Number of hopsital admissions per year | 5-11 | 12-17 | 18+ |
| --- | --- | --- | --- |
| 1 | 81.4256 | 79.9087 | 85.0762 |
| 2 | 12.2001 | 9.58904 | 8.91194 |
| 3 | 3.28992 | 3.50076 | 2.37087 |
| 4 | 1.16518 | 2.43531 | 0.973751 |
| 5 | 0.891021 | 0.913242 | 0.402202 |
| 6 | 0.0027416 | 0.00608828 | 0.00529213 |
| 7 | 0 | 0.456621 | 0.275191 |
| 8 | 0.13708 | 0.152207 | 0.127011 |
| 9 | 0.0685401 | 0 | 0.169348 |
| 10+ | 0.548321 | 2.43531 | 1.16427 |
